# Nasal DNA methylation at three CpG sites predicts childhood allergic disease

**DOI:** 10.1101/2022.06.17.22276520

**Authors:** Merlijn van Breugel, Cancan Qi, Zhongli Xu, Casper-Emil Tingskov Pedersen, Ilya Petoukhov, Judith M. Vonk, Ulrike Gehring, Marijn Berg, Marnix Bügel, Orestes A. Capraij, Erick Forno, Andréanne Morin, Anders Ulrik Eliasen, Yale Jiang, Maarten van den Berge, Martijn C. Nawijn, Yang Li, Wei Chen, Louis Bont, Klaus Bønnelykke, Juan C. Celedón, Gerard H. Koppelman, Cheng-Jian Xu

## Abstract

Childhood allergic diseases, including asthma, rhinitis and eczema, are prevalent conditions that share strong genetic and environmental components. Diagnosis relies on clinical history and measurements of allergen-specific IgE. We hypothesized that a multi-omics model could accurately diagnose childhood allergic disease. We show that nasal DNA methylation has by far the strongest predictive power to diagnose childhood allergy, surpassing blood DNA methylation, genetic risk scores, and environmental factors. DNA methylation at only three nasal CpG sites classifies allergic disease in Dutch children, with an area under the curve (AUC) of 0.86. This was replicated in US Hispanic children (AUC 0.82). DNA methylation at these CpGs additionally detects allergic multimorbidity and symptomatic IgE sensitization. Using nasal single-cell RNA-sequencing data, we map these three CpG sites to reflect the influx of T cells and macrophages that contribute to allergic inflammation. Our study offers a simple, non-invasive diagnostic test for childhood allergy.

## Introduction

Allergic diseases such as asthma, rhinitis, and eczema are highly prevalent, non-communicable childhood diseases worldwide^1^ that have a significant impact on quality of life and cause a considerable burden on healthcare systems^2,3^. Allergic diseases often coexist in the same individual, suggesting shared underlying mechanisms^4^. The prevalence of allergic diseases has increased rapidly for more than 50 years, and 40-50% of schoolchildren in the Western world have allergen-specific Immunoglobulin (IgE) to one or more common allergens^5^. However, the presence of specific IgE does not necessarily coincide with presenting allergy symptoms, and effective biomarkers are needed to capture the allergic inflammation signal rather than the presence of IgE. There is therefore a great need for non-invasive biomarkers to facilitate better diagnosis, especially in early childhood.

Childhood allergic diseases have strong shared genetic and environmental components^6,7^. Many shared genetic risk variants for allergic disease influence the expression of immune-related genes^8^. The increase in prevalence of allergic diseases indicates that environmental exposures, such as microbial stimulation^9^, air pollution, breastfeeding, pets at home, and cigarette smoking, also play an important role^10^. Environmental factors can have sustained effects on gene expression through the modulation of epigenetic features such as DNA methylation^11^. Indeed, epigenome-wide association studies (EWAS) of allergic diseases in both whole blood and nasal-brushed cells have revealed many shared epigenetic signatures of allergy^12–15^.

There is a need for the prediction of individual allergic disease risk, especially in young, preschool children, in whom allergy is difficult to diagnose. While previous studies have constructed models to assess individual disease risk, these models investigated usually only a single layer of ‘omics’ data, such as the genome, epigenome, transcriptome, proteome, or metabolome^14,16–19^. Given the complex biological nature of allergic diseases, with the known involvement of multiple omics layers and environmental factors, leveraging the information from multi-omics data may improve prediction of these diseases^20^. Recently, an integrated multi-omics approach was successfully used to predict cytokine production of immune cells^21^, but there are few complex disease prediction models using multi-omics approaches.

In this study, we developed a machine learning prediction model for allergic disease, by systematically integrating large-scale multi-omics data from the genome, DNA-methylome of blood and nasal cells, and environmental factors. We first evaluated the predictive power of each layer separately and identified the most predictive features to construct a parsimonious allergy prediction model in the Dutch PIAMA (Prevention and Incidence of Asthma and Mite Allergy) birth cohort^22^. We replicated our model in three independent birth cohorts of children of different ethnicities and ages. We report that allergy can be predicted accurately using nasal DNA methylation at only three CpG sites. These sites also detect allergic multimorbidity and symptomatic sensitization, and can be mapped to influx of T cells and macrophages in the nasal mucosa.

## Results

### Nasal DNA methylation has most predictive power for allergic disease

To construct a multi-omics prediction model of allergic disease, we first investigated the relative contributions of each data layer, including genomics, blood and nasal DNA methylation, and personal and environmental factors. We performed these analyses in the PIAMA cohort, a prospective national population-based birth cohort study on the development of asthma and allergy in the Netherlands^22^ (Figure 1-a). Here, allergic disease was defined by both the presence of asthma/rhinitis/eczema and the detection of allergen specific-IgE in blood; we found a prevalence of 19.3%.

**Figure 1.**
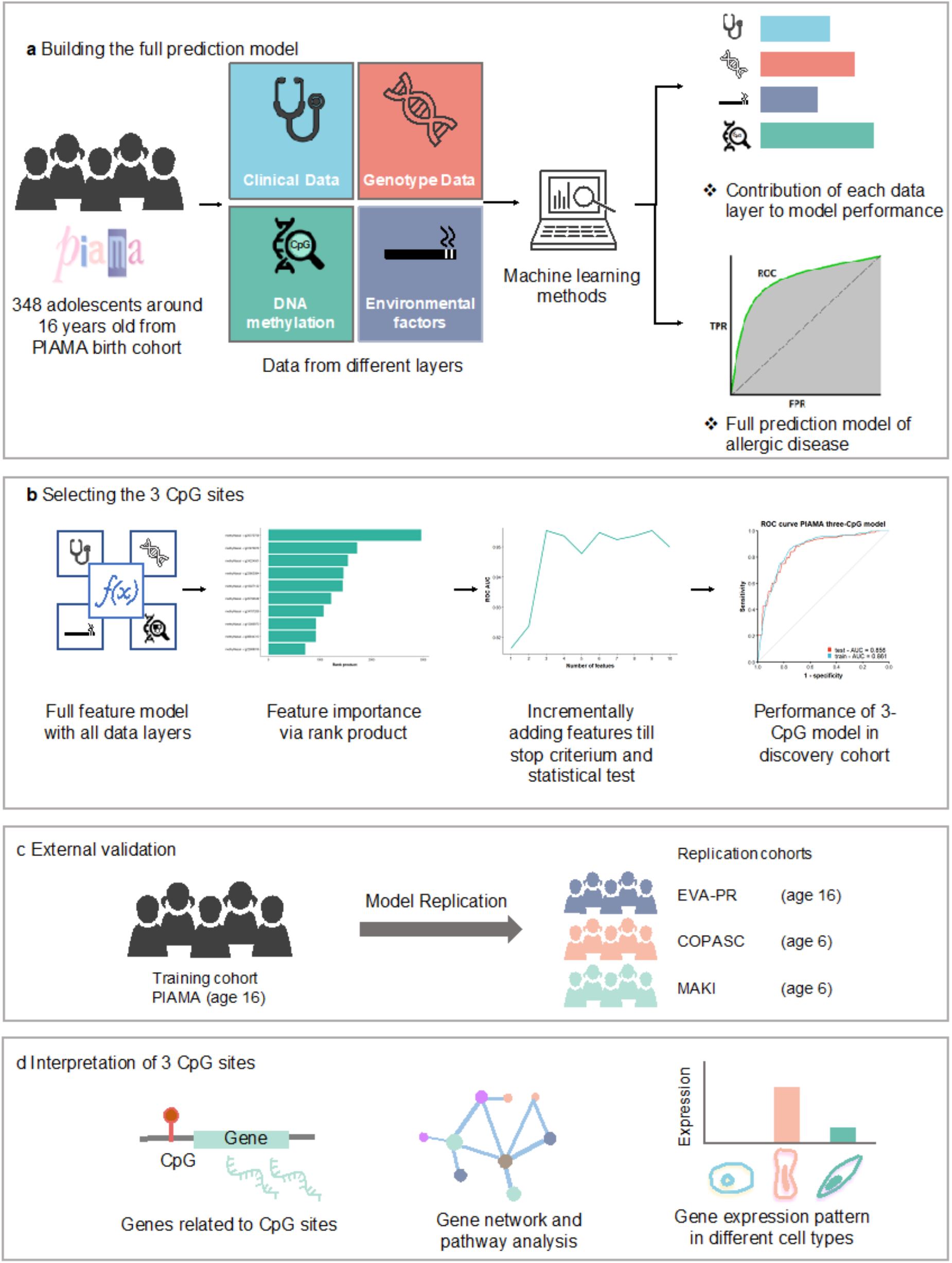
Study design. (a) We integrated multi-omics data including, genetics, DNA methylation from blood and nasal brushes, and environmental factors, from the PIAMA cohort. Use machine learning methods, we aimed to 1) assess the contribution of each data layer to the performance of a prediction model; and 2) present a simple prediction model for allergic disease. (b) To develop a parsimonious prediction model, we first ranked all features from the full dataset, then features were added incrementally until no significant increase in performance was seen. The performance of the final parsimonious model was demonstrated in the discovery cohort. (c) The final model was evaluated by applying it to another similar but independent cohort (EVA-PR) and two independent younger children cohorts, COPSAC2010 and MAKI. (d) To understand the information that was captured by the three CpG sites used in the model, we linked the methylation level of the CpG sites to the expression of genes by eQTM analysis. The eQTM genes were functionally interpreted by gene network and pathway analysis; scRNA-seq data were used to interpret the expression pattern of eQTM genes in different nasal cell types.

In 348 children aged 16 years with complete data (Table 1), we selected features from four data layers (genome, blood and nasal methylome, environmental factors) that had been previously associated with allergic disease (see Methods). In total, we analyzed 467 features. We evaluated six machine learning methods (see Methods) and adopted Elastic Net^23^ as our model of choice because it offered accurate performance, low overfit, and good interpretability (Table S1).

**Table 1.** Characteristics of the discovery and replication cohorts.

|  | **PIAMA**<br>**(Discovery)** | **EVA-PR**<br>**(Replication)** | **COPSAC2010**<br>**(Replication)** | **MAKI**<br>**(Replication)** |
| --- | --- | --- | --- | --- |
| Sample size | 348 | 481 | 503 | 259 |
| Age (years) | 16.3 (0.20) | 15.5 (3.0) | 6.0 (0.2) | 5.8 (0.4) |
| Gender male | 179/348 (51.4%) | 248/481 (51.6%) | 217/503 (43.1%) | 146/259 (56.4%) |
| Smoking | 37/297 (12.5%) | NA | 35/ 503 (6.9%) | 15/259 (5.8%) |
| Asthma | 23/348 (6.6%) | 236/481 (48.9%) | 31/503 (6.2%) | 20/255 (7.8%) |
| Rhinitis | 59/348 (17.0%) | 295/481 (61.3%) | 60/503 (53.3%) | 27/259(10.4%) |
| Eczema | 29/348 (8.3%) | 89/481 (18.5%) | 55/503 (10.8%) | 28/259 (10.8%) |
| Positive allergen-specific IgE | 162/348 (46.6%) | 311/481 (64.7%) | 85/503 (18.7%) | 56/ 205 (27.3%) |
| Allergy | 67/348 (19.3%) | 267/481 (55.5%) | 37/503 (8.9%) | 29/259 (11.2%) |

Continuous variables are presented as mean (SD); categorical variables are presented as (number of "yes")/(total number (with this phenotype available)) (percentage).
PIAMA Prevention and Incidence of Asthma and Mite Allergy birth cohort (Netherlands); EVA-PR Epigenetic Variation and Childhood Asthma in Puerto Ricans (USA); COPSAC2010 Copenhagen Prospective Study on Asthma in Childhood (Denmark); MAKI trial (the Netherlands)

The largest predictive power was attributed to DNA methylation levels in nasal epithelial cells (Figure 2-a); other layers had a negligible effect. This conclusion did not change when we varied the order in which the layers were added. 86% of the top-50 features were nasal and 14% were blood DNA methylation CpG sites (Table S2), underlining the predictive power of the DNA-methylome. These results indicate that genetic factors had very limited predictive power for allergic disease at the individual level, which was also observed in a larger dataset of 675 individuals from the PIAMA cohort for whom both genotype and allergy phenotype data were available (Figure S1).

**Figure 2.**
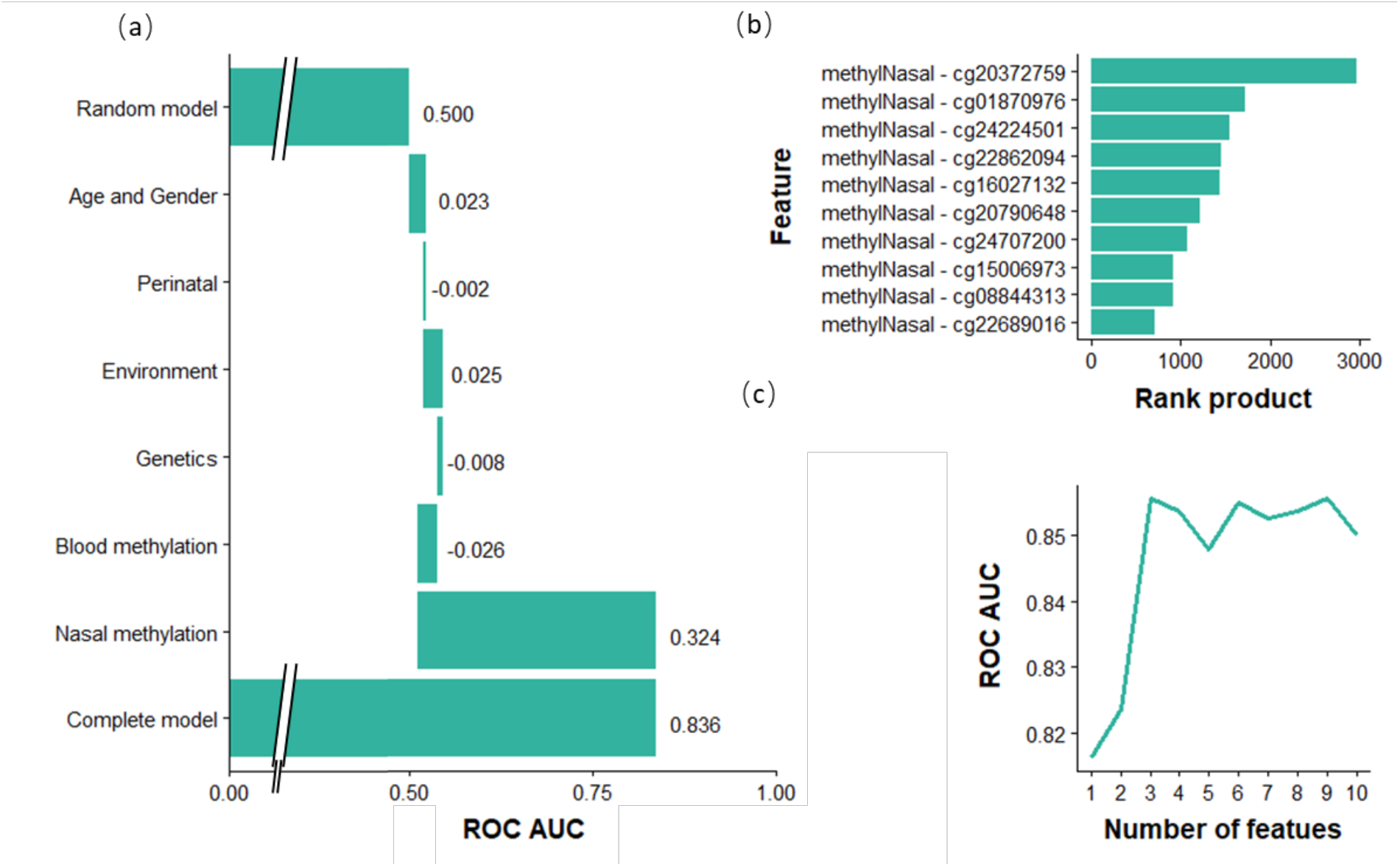
Feature selection. (a) Based on validation performance of an Elastic Net model using a 10-times repeated 10-fold cross-validation procedure. Layers were added sequentially: age and gender were added first, then perinatal factors were included in the model, etc. Negative contributions could occur when variables had low predictive power and increased model overfit. Perinatal features were low birth weight and breastfeeding; Environment: pets during pregnancy, maternal smoking and older siblings at home; Genetics: allergy SNPs and polygenic risk scores (PRS) for the combined allergy phenotype, as well as for asthma, rhinitis, eczema and IgE sensitization; Blood methylation: 219 CpG sites from blood cells previously associated allergy; Nasal methylation: 134 CpG sites from nasal cells previously associated with allergy. (b) Rank product of top-10 features. (c) The AUC of models with an incremental number of features; after the first three nasal CpG sites, adding another CpG site did not further increase the model’s performance.

### A parsimonious prediction model with only three nasal CpG sites predicts allergy

We next aimed to generate a parsimonious model to increase our model’s robustness, reproducibility, and cost-effectiveness, and to facilitate translation to a clinical setting (Figure 1-b), starting with the top-10 ranked features (Figure 2-b), all of which were nasal DNA CpG methylation sites (Table S3). When we compared the performance of models with an incremental number of features, we observed that after including the first three nasal CpG sites, additional CpG sites did not further increase model performance (p-value = 0.76, corrected repeated k-fold cross-validation (cv) test^24^, Figure 2-c). These 3 CpGs (cg20372759, cg01870976 and cg24224501) are located near gene *CYP27B1, SNRPA1* and *LRRC17* respectively (Table S4). The ‘3-CpG sites’ model performed well in predicting allergic disease in the PIAMA discovery cohort, with an ROC AUC (receiver operating characteristic, area under the curve) test score of 0.86 (Figure 3-a). This parsimonious model has near-zero overfit (ROC AUC difference of 0.005), as the ROC AUC training score was also 0.86 (Figure 3-a). Because of the imbalance of cases and controls in the discovery dataset, we also introduced the precision-recall curve (PRC)^25^ to evaluate model performance: the PRC AUC of the model was 0.50 (Figure S2-a). The model’s coefficients can be found in Table S5. The 3-CpG model was also compared to the previously published nasal 30-CpG model that performed well in predicting atopy (i.e. the presence of IgE sensitization) in the Epigenetic Variation and Childhood Asthma in Puerto Ricans study (EVA-PR)^14^. Although we only used three nasal CpG sites, our model showed a comparable performance (p-value = 0.32, corrected repeated k-fold cv test) with the more complex 30-CpG model (ROC AUC test score of 0.83) (Figure S3-a). Overfit of this more complex model remained low (ROC AUC difference of 0.008), albeit slightly larger than our 3-CpG model. The PRC AUC of 0.48 was also lower in the 30-CpG model (Figure S3-b), although the difference was not significant (p-value = 0.40, corrected repeated k-fold cv test).

**Figure 3.**
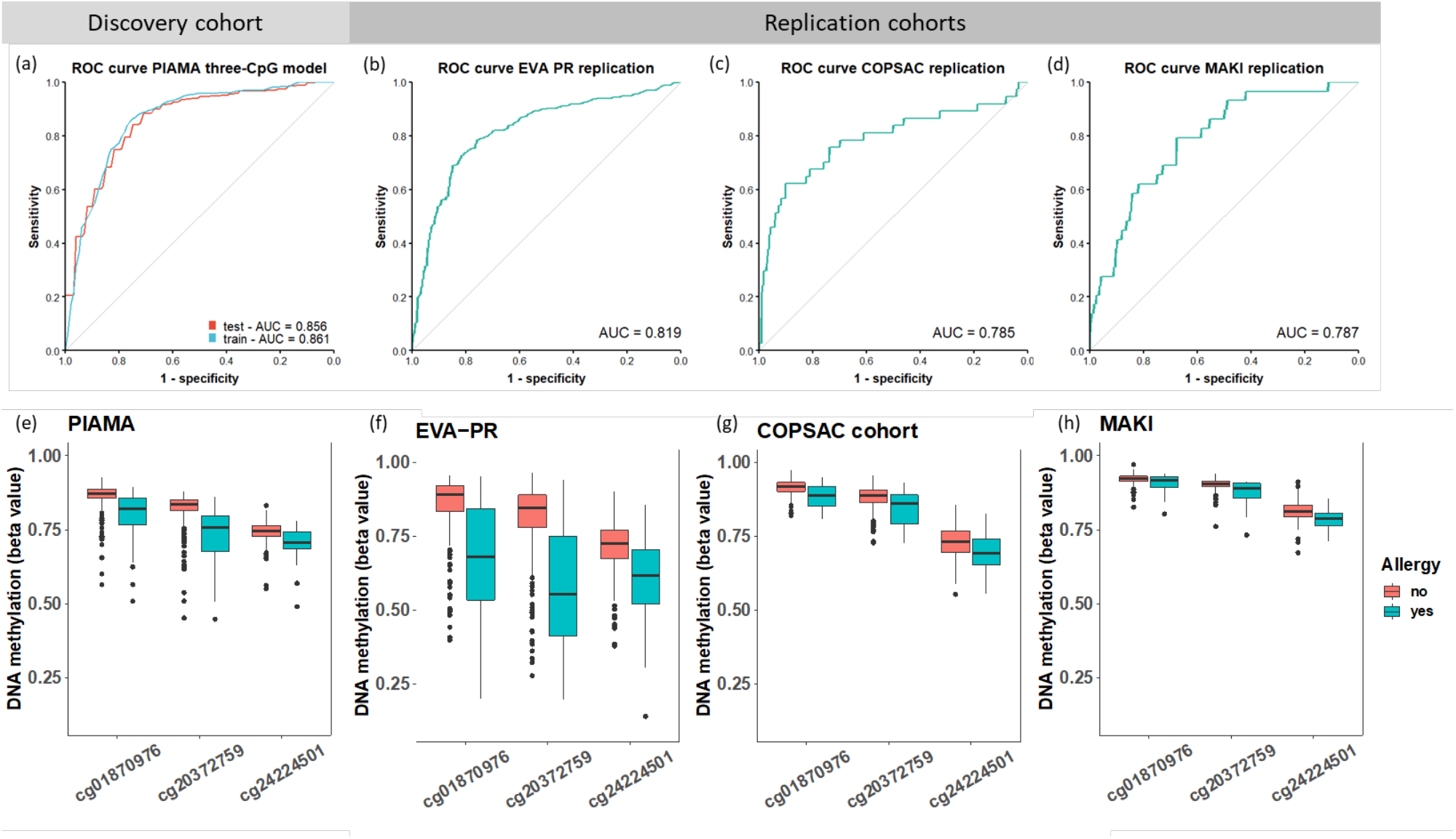
Model performance. The ROC curve and AUC of the model in the discovery cohort (a) and replication cohorts: EVA-PR (d, mean age 16 years), COPSAC2010 (e, mean age 6 years) and MAKI (f, mean age 6 years). The DNA methylation levels of the three CpG sites in subjects with and without allergy are shown in e-h, for PIAMA, EVA-PR, COPSAC2010 and MAKI, respectively.

### Three-CpG model also predicts allergic disease in US Hispanic children

To test its generalizability across ethnicities, we first replicated our 3-CpG model in a cohort of comparably aged US Hispanic children from Puerto Rico (EVA-PR)^14^ (Figure 1c). In EVA-PR, the prevalence of allergic disease was 55.5% and the mean age was 15.5 years (Table 1). Our model performed well on both ROC AUC (0.82) (Figure 3-b) and PRC AUC (0.84) (Figure S2-b). The PRC AUC’s baseline value (of a random model) is equal to the disease prevalence, here 0.56, which explains its steep increase relative to the discovery cohort. The good replicability is affirmed by the precision and recall metrics of 0.80 and 0.79, respectively, for the EVA-PR cohort, compared to 0.53 and 0.64 in the PIAMA cohort (Table S6).

We next extrapolated our model to two cohorts of younger children (6-year-olds; COPSAC2010, Denmark and MAKI trial, the Netherlands, Table 1), both with an ROC AUC of 0.79 (Figure 3-c, d). Our model’s performance in the younger cohorts showed considerably lower recall (Figure S2-c, d), especially in MAKI where recall dropped to 0.03 (Table S6). This indicates we should be cautious in extrapolating our prediction model to this younger age group. In order to understand the difference in performance between age groups, we compared the probe distribution of the three CpG sites in subjects with and without allergic disease in all four discovery and replication cohorts (Figure 3-e-h). We observed that the DNA methylation levels of the three CpG sites were clearly lower in subjects with allergic disease than in subjects without allergic disease in all cohorts, with the highest difference seen in adolescents and paralleled by the better prediction power in these two cohorts. The DNA methylation levels in control samples were higher in the younger cohorts compared to the adolescent cohorts, which may be attributed to the ageing effect on DNA methylation (Figure 3-e-h).

### DNA methylation at three CpG sites relates to symptomatic IgE sensitization and disease multi-morbidity

Allergic disease was defined as a combination of diseases (asthma/rhinitis/eczema) and IgE sensitization. To better understand the association of the three top CpG sites with both disease and IgE sensitization, we performed group by group comparisons in the discovery cohort, analyzing symptomatic and asymptomatic children with and without IgE sensitization. The results showed that these CpG sites were not only associated with IgE sensitization, but also with the presence of clinical symptoms in IgE sensitized subjects (Figure 4-a). Importantly, we observed significant differences between the IgE+ symptom+ group and the IgE+ symptom-group for the three CpG sites (p-values = 1.4×10^−3^, 6.2×10^−4^ and 9.8×10^−4^, for CpG cg01870976, cg20372759, cg24224501, respectively, two-sided Student’s t-test), suggesting that these three sites distinguish symptomatic from asymptomatic sensitization. Next, we found lower DNA methylation of these CpG sites in multimorbidity compared to a single allergic disease (p-values = 0.039, 0.047 and 0.17, two-sided Student’s t-test, Figure 4-b). These findings were also exactly replicated in the EVA-PR cohort (Figure 4-c, d), confirming that nasal DNA methylation captures differences between asymptomatic and symptomatic sensitization and allergic disease multimorbidity.

**Figure 4.**
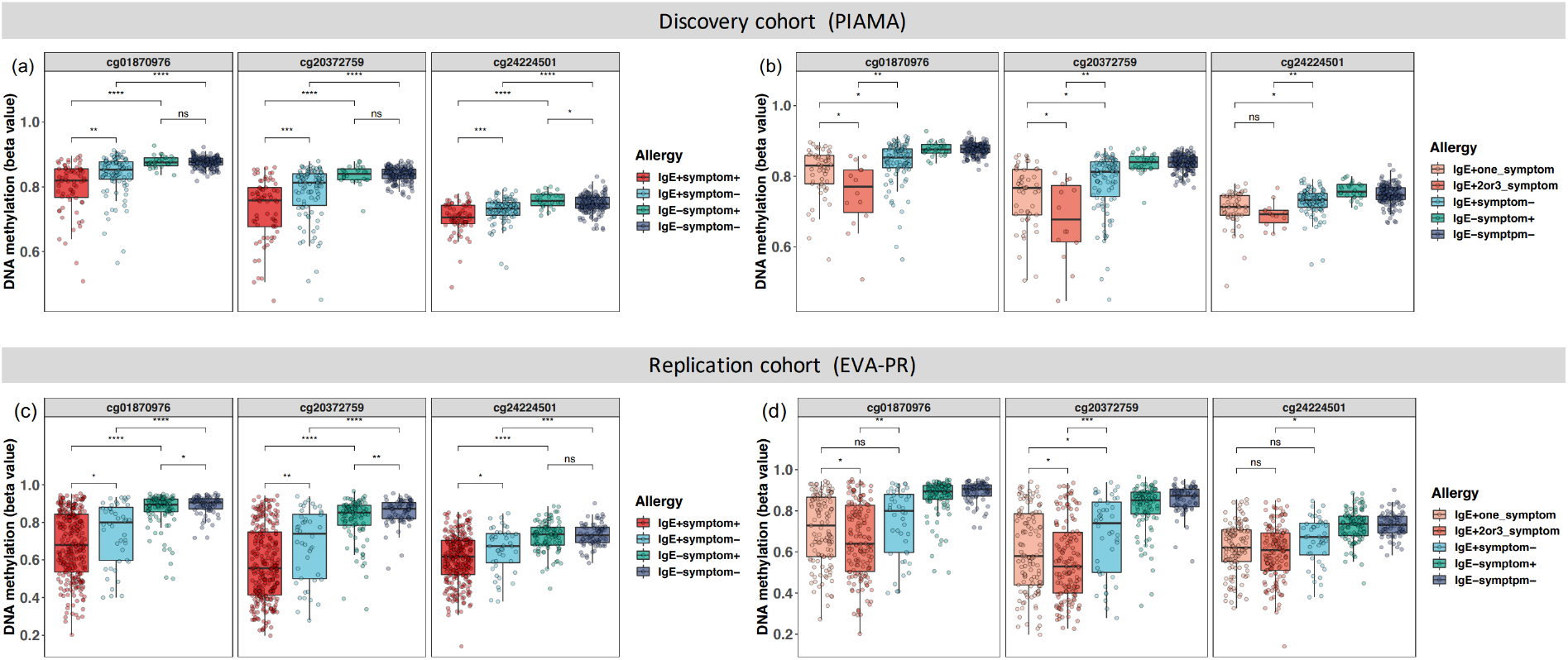
DNA methylation levels of three CpG sites in subgroups of allergy phenotype in the discovery (a, b, PIAMA) and a replication cohort (c, d, EVA-PR). Samples were stratified by both IgE sensitization and symptom (a, c), and were additionally stratified by number of symptoms (b, d). The Student’s two-sided t-test was used to compare the difference between different groups (* P<0.05; ** P<0.01, *** P<0.001, **** P<0.0001, ns not significant).

### Genes associated with the three CpG sites identified a signature of immune cells

To interpret the biological changes responsible for the differential methylation levels of the three nasal CpG sites in allergy (Table S4), we first correlated DNA methylation to gene expression levels by expression quantitative trait methylation (eQTM) analysis, using matched nasal RNA-seq data from children of the PIAMA cohort (n=244 with both types of data available). Expression levels of 127 unique genes (eQTM genes) were associated with methylation levels at the three CpG sites, resulting in 182 CpG-gene pairs (Table S7), with many genes related to more than one of the three CpG sites (Figure S4). We applied weighted correlation network analysis (WGCNA^26^) on these eQTM genes and identified two modules (Table S8). Using KEGG pathway enrichment analysis, we found that genes from Module 1 were enriched in ten, mostly T-cell related pathways, including hematopoietic cell lineage, Th1 and Th2 cell differentiation, and Th17 cell differentiation (Figure S5-a). Genes from Module 2 were enriched in eight pathways (Figure S5-b), including the B cell receptor signaling pathway and complement and coagulation cascades. These results suggest that these eQTM genes are related to immune cells and may be involved in the allergic inflammatory response.

We hypothesized that the differences in CpG methylation in nasal brushes in allergic and non-allergic subjects reflected differences in cell type composition or different DNA methylation profiles in different cell types, with the eQTM genes reflecting the cell-type specific transcriptional profile. To map these putative cell types, we generated a single-cell RNA-seq dataset from nasal brushings of 9 adult subjects (5 healthy and 4 asthmatics, Table S9) to assess the cell-type specific expression pattern of the eQTM genes. We identified 10 cell types: 8 epithelial cell clusters (basal, goblet 1, goblet 2, squamous, cycling, ciliated, deuterosomal, and ionocytes), and 2 immune cell clusters (myeloid and T cells) (Figure 5-a). Most of the eQTM genes from Module 1 were highly expressed in the T cell cluster, while the genes from module 2 were enriched in the myeloid cell cluster (Figure 5-b). Subsetting and re-clustering of all immune cells (Figure 5-c) revealed the presence of 5 myeloid cell clusters (dendritic cells (DC) -1, DC-2, macrophages, classical monocytes and plasmacytoid dendritic cell (pDCs), Figure 5-e), 3 T cell clusters (CD8 T cells/ cytotoxic T lymphocytes (CTLs), CD4 cells/ Treg cells, and NK cells, Figure 5-f), and single cluster representing B cells, plasma cells and mast cells. Genes from Module 1 were most highly expressed in CD8/ CTLs and CD4/ Treg cells (Figure 5-d, h), while genes from Module 2 were enriched in macrophages (Figure 5-d, f), suggesting that the three CpG sites may capture the relative increase in both T-cells and macrophages in allergic inflammation in nasal brushes.

**Figure 5.**
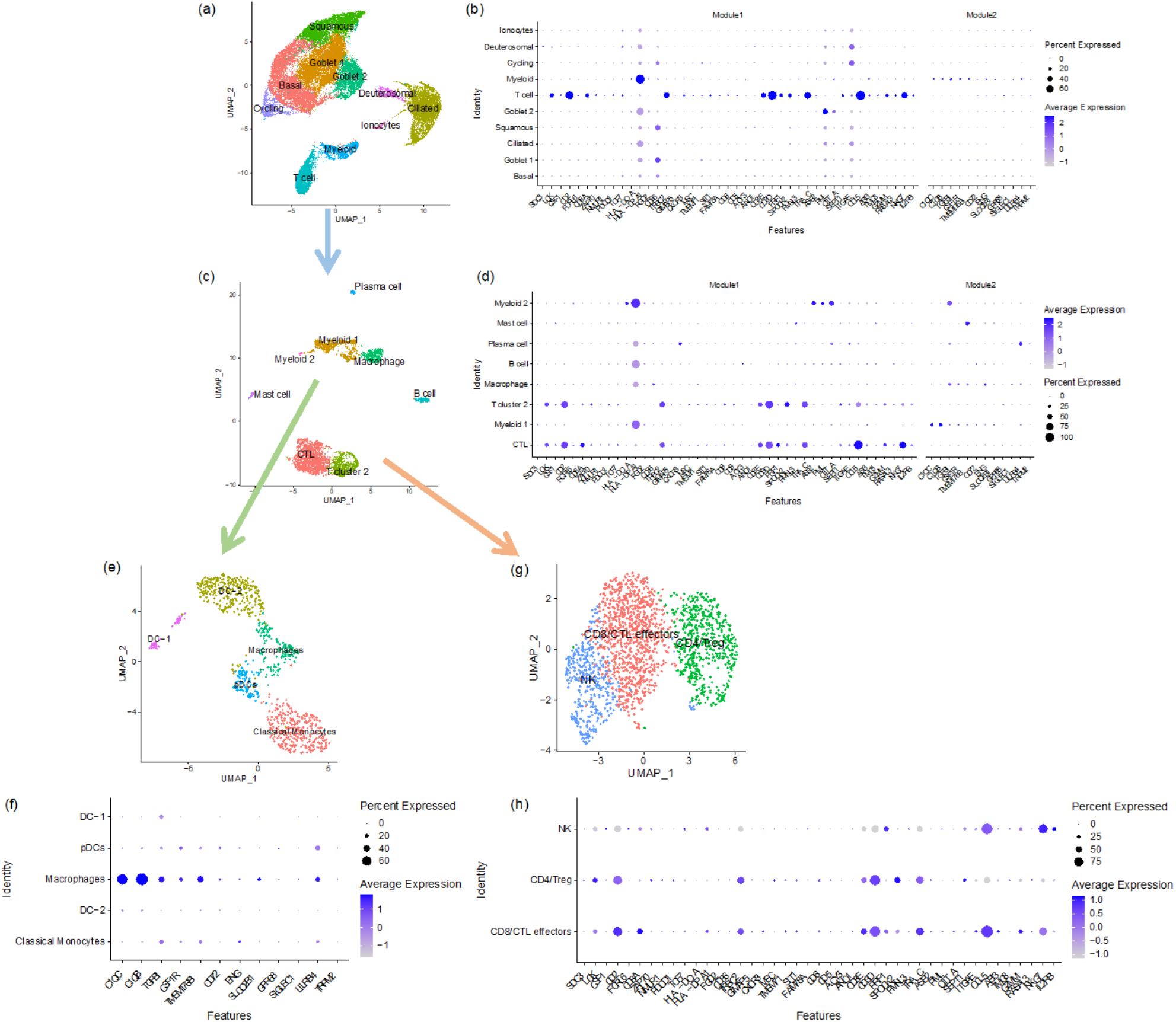
Expression patterns of genes associated with three CpG sites in different cell types. Using single cell data, we identified 10 cell types in nasal brushing cells (a). We identified two gene modules from eQTM genes by Weighted Gene Co-expression Network Analysis (WGCNA). Plots (b) depicts the average expression levels per cell cluster of genes from Module 1 and Module 2 which were available in the nasal scRNA-seq dataset in all cell types. Genes from Module 1 were highly expressed in T cell cluster and genes from Module 2 were enriched to myeloid cells. After re-clustering of all immune cells from the two immune clusters, we identified seven immune cell clusters (c). Plots (d) showing the genes from Module 1 was enriched to T cell clusters and genes from Module 2 was enriched to myeloid cell clusters. Further re-clustering of T cell cluster (f) and myeloid cell cluster (e) respectively identified that genes from Module 1 were enriched to CD8/CTL effectors and CD4/ Treg cells (h), while genes from Module 2 were enriched to macrophages (f). CTL: Cytotoxic T Lymphocyte; DC: dendritic cell; pDCs: Plasmacytoid dendritic cells; NK: nature killer cell; Treg: regulatory T cell.

### Allergy-associated SNP may mediate allergy through DNA methylation of a nasal CpG site

We performed methylation quantitative trait locus (MeQTL) analysis to investigate if the three nasal CpG sites are regulated by allergy-associated SNPs. For this, we associated the three CpG sites with SNPs previously associated with allergic disease^8^. We identified 15 SNP-CpG pairs showing nominal significance (p-value <0.05, Table S10), suggesting potential regulation of genetic variants at the methylation level of these CpG sites. The most significant SNP-CpG pair (rs9372120-cg20372759) was also associated with allergic disease in our PIAMA dataset. We assessed the mediation effect of this SNP, located in the *ATG5* (*Autophagy Related 5*) gene, on allergic disease through DNA methylation and found that 75.7% of the effect of rs9372120 on allergic disease was mediated by DNA methylation of cg20372759 (Table S11).

## Discussion

In this study, we integrated multi-omics data to assess the ability of a combination of genetic, epigenetic and environmental factors to predict allergic disease. We hypothesized that multi-omics integration would provide a more accurate allergy prediction model, compared to using a single omics layer. However, our study has shown that nasal DNA methylation alone is a strong and valuable biomarker for diagnosing allergic disease. Thus, we provide a proof-of-concept that DNA methylation can be used to generate a simple diagnostic test for a complex disease such as allergy. We extended this finding by showing that nasal DNA methylation is able to differentiate symptomatic from asymptomatic IgE-sensitization, providing for the first time a set of biomarkers that can distinguish these two conditions, as well as reflecting allergic disease comorbidity. We mapped the changes in these three CpG sites to an increase in T cells and macrophages in the nasal mucosa. We propose that nasal DNA methylation accurately reflects allergic inflammation in the nasal mucosa, driven by activated T cells and macrophages.

Allergic disease often starts at an early age and may manifest in different organs, such as the respiratory system (asthma, rhinoconjunctivitis), the skin (atopic eczema), and the gastrointestinal tract (food allergy). These allergic diseases may be present at the same time in an individual (comorbidity) or may have a temporal sequence, such as displayed in the allergic march (from eczema to asthma and hay fever)^4,27^. Unsupervised statistical techniques performed in large population-based cohorts suggested that two main clusters could be observed: unaffected children and children with allergic comorbidity^28^. This pattern of extensive comorbidity of allergic diseases was further supported by the identification of genetic and epigenetic variations that were associated with all three diseases (asthma, rhinitis and eczema)^8,29^. Part, but not all, of this overlap is explained by IgE sensitization^4^, suggesting that there are more pathogenic mechanisms shared between allergic diseases. These findings prompted us to investigate allergic disease as one unified phenotype.

One of our key findings is that DNA methylation of nasal brushings is most predictive for allergic disease. This can be explained by the fact that epigenetic signatures in the upper airways may reflect both genetic and environmental factors, in addition to cell-type composition and cell activation^30^. The three CpG sites that we use in our parsimonious model are related to genes that were enriched in immune cell pathways and mapped to T cells and macrophages. However, a limitation of our single-cell annotation is the lack of eosinophils in the nasal single-cell RNA-seq dataset, as these cells are selectively missed due to their high RNase content. Our findings indicate that the influx of immune cells into the nasal mucosa can be detected using nasal brushes and provides a strong and distinctive DNA methylation signal in allergic disease. In addition, we found that DNA methylation might mediate the effect of genetic variants on allergic disease. Taken together, these results indicate that our CpG sites may capture both genetic contribution and inflammation in the nose.

In contrast to the nasal brushings, methylation levels of CpG sites in blood did not improve the performance of our prediction model. One explanation might be that the signal in blood is diluted due to the complex mixture of immune cells present in the sample. We previously reported a similar observation in childhood asthma, showing that the DNA methylation signal associated with asthma was stronger in purified eosinophils than in whole blood samples^12,31^. Another explanation could be that the tissue-resident T cells and macrophages may be different from the counterparts in blood which do not have the same methylation levels at these three CpG sites. In support of this, methylation at the three CpG sites in blood showed no obvious differences between subjects with and without allergy (Figure S6).

Along with making an accurate prediction model, we also aimed to better understand the role of the selected CpG sites in allergic disease. In this study, allergic disease phenotype was defined by the combination of IgE sensitization and parent-reported symptoms following the definitions of the Mechanisms of the Development of ALLergy (MeDALL) consortium^4^, which takes both IgE-mediated mechanisms and multimorbidity into consideration. By creating subsets of subjects based on phenotype status, we could examine the role of the three CpG sites on IgE sensitization and multimorbidity. Although IgE sensitization is generally used to define the allergic status, sensitization to allergens does not necessarily mean the subjects have allergy symptoms^32^. Asymptomatic allergic subjects may differ from symptomatic subjects in total serum IgE levels, mono- or polysensitization, presence of Treg cells, and basophil reactivity^33^. Therefore, we urgently need effective biomarkers that can distinguish asymptomatic sensitization from full-blown allergic disease. In our study, the nasal DNA methylation levels of three CpG sites were lower in symptomatic than asymptomatic IgE-sensitized subjects, showing the potential of these three CpG sites in nasal epithelial cells to be a biomarker for the presence of symptoms in IgE-sensitized individuals, or in other words, to be a more selective biomarker for allergic disease. Thus, these CpG sites are not simply a nasal reflection of the presence of specific IgE, but they also capture the presence of symptomatic disease. In addition, the association of methylation levels at these three CpG sites and the number of allergic symptoms (asthma/ rhinitis/ eczema) strongly suggest that our findings could also be used as a biomarker for allergic multimorbidity, adding more clinical relevance to our work. These findings were strongly replicated in a population of Hispanic children, showing that our prediction model is valid across at least two ethnicities.

Contrary to the predictive power of DNA methylation, genetics did not contribute to the performance of our model. At first sight, inherent genetic variants show great potential for early diagnosis of various diseases, including obesity, coronary artery disease, and Alzheimer’s disease^16,34,35^. Large genome-wide association studies (GWAS) have identified many genetic variants associated with allergic disease, which enables allergic disease to be predicted based on polygenetic risk scores. However, significant genetic variants identified by a recent GWAS only explained a small proportion of disease heritability (3.2% for asthma, 3.8% for hay fever, and 1.2% for eczema)^8^, and these identified variants were mostly common variants with relatively small risk effects and ones that tend to be difficult for correctly identifying risk at the individual level, thus offering limited predictive power for allergic disease^36^. Dijk et al^37^ generated a prediction model for asthma during the first eight years of life based on environmental factors and genetic risk scores in 1858 children of the PIAMA study. Genetic risk scores based on SNPs identified by a large asthma GWAS did not add predictive power for asthma over using just family history and environmental factors. This was also shown in an independent Swedish cohort study.

While our three-CpG model can predict the presence of allergy, several limitations should be taken into consideration. Firstly, our model was trained in Dutch adolescents with a narrow age range. For the two cohorts of younger children (COPSAC2010 and MAKI), model extrapolation resulted in a low recall rate, in contrast to the good replicability in the independent adolescent cohort. This may be explained by changes of DNA methylation with aging^38^. Therefore, extra caution is needed when extending our model to younger children and further studies should address if these, or other CpG sites, can contribute to diagnosing allergy in younger children. Secondly, as our model was trained using cross-sectional data, we could not assess if our model could predict future disease status. Finally, our findings that genetic variants did not contribute to the prediction of allergic disease might be due to our relatively small dataset. However, it has been reported that also in larger datasets, genetic risk variants of allergic sensitization had limited value for clinical prediction at an individual level^39^.

This study has three implications for biomarker discovery. Firstly, on a general level, the combination of nasal methylation sites is suggested to be a stable biomarker^40^ that is replicable across different ethnicities. Secondly, there are promising implications for the field of pediatric allergy: physicians are looking for accurate non-invasive methods to diagnose diseases, especially in young children, and to reduce misdiagnoses^41^. Nasal DNA methylation biomarkers would only require a simple nasal swab rather than a blood test to assist physicians in making a diagnosis. In addition, our model’s parsimonious nature – with only three CpGs – makes it more cost-effective and shows the potential to distinguish symptomatic and asymptomatic sensitization and disease comorbidity. Thirdly, epigenetics biomarkers hold encouraging implications for the future. Besides diagnosis, they can also be used to facilitate decisions on personalized medicine and treatment selection. In cancer research, this has already proved to be feasible and valuable^42^.

In conclusion, we have demonstrated that nasal DNA methylation has good prediction power for allergic diseases. We have presented a parsimonious model using only three nasal CpG sites that can predict allergic disease in adolescents; it was successfully replicated in an independent adolescent cohort. We show that nasal methylation bears information on the presence of allergic disease in the presence of IgE sensitization, and also reflects allergic disease multimorbidity. These CpG sites reflect the influx of T cells and macrophages that contribute to allergic inflammation.

## Methods

### Study and population description

The discovery analysis was performed in the PIAMA birth cohort. Details of the cohort have been published previously^22^. The current study used data from the 16-years follow-up, and included validated questionnaires, blood testing for IgE sensitization, and DNA isolation of whole blood cells and nasal brushes. In this study, an allergic disease case was defined as a child with at least one of three allergic diseases (asthma/rhinitis/eczema) in combination with the presence of IgE for at least one allergen. Details on allergic disease definitions, specific IgE measurements, and study questionnaires can be found in the online supplementary materials. In total, we had complete data for all layers from 348 individuals. Characteristics of the dataset used in this analysis compared to the initial PIAMA dataset can be found in Table S12. Detailed numbers of subjects with each disease and IgE sensitization in PIAMA can be found in Figure S7.

### Data measurement and quality control

#### Genotype data

DNA was extracted from whole blood samples and nasal brushing samples which were collected from the lower inferior turbinate. Genotyping was performed in four phases using four different platforms, being Illumina Human610 quad array, Illumina HumanOmniExpress array, Illumina Human Omni Express Exome Array, and Illumina Infinium Global Screening Array. Quality control (QC) of each phase was performed and then the data were merged together. SNPs were harmonized by base pair position annotated to genome build 37. In total, 2075 individuals remained after quality control, and their data from the four platforms were merged together. Then imputation was performed using the Michigan server with a reference panel of HRC.r1. SNPs of high quality (imputation quality score Rsq >0.8), MAF>0.01 and HWE <1×10^−12^ were used for further analysis. After stringent QC, 2075 samples and 2,893,496 SNPs remained.

#### DNA methylation data

Blood and nasal DNA methylation were measured by Infinium HumanMethylation450 BeadChip array. Detailed QC steps were published previously^29^. After QC, 613 blood samples and 478 nasal samples with 436,824 CpG probes remained. Methylation β-values at a given CpG were derived from the ratio of the methylated probe intensity to overall intensity (sum of methylated and unmethylated probe intensities). Then β-values were transformed to methylation values (M-values) as log_2_(β/(1-β)), which were used in the downstream analysis.

#### RNA-seq data

Total RNA was also extracted from nasal brushing samples and was sequenced by Illumina HiSeq2500 sequencer using default parameters for paired-end sequencing (2 × 100 bp).

Detailed QC and alignment steps were reported previously^29^. In total, 17,156 expressed features and 326 samples passed QC. After matching with DNA methylation data, 244 samples remained for downstream analysis. Raw count data were transformed to log_2_CPM using the *voom* function in the R package *limma*^43^.

### Modeling approach

#### Feature selection

For feature selection, we selected an initial list of promising candidate variables based on previous research. The features included:

##### Environmental factors

perinatal/environmental factors collected using questionnaires during pregnancy and the first years of life (pets during pregnancy, maternal smoking, breastfeeding, older siblings at home, and low birth weight^44,45^).

##### Genetic factors

136 SNPs were chosen based on their strong association with the allergy phenotype in a GWAS^8^, of which 75 passed our QC thresholds. Moreover, the polygenic risk score (PRS) for allergy was calculated from 4813 allergy-associated SNPs^8^, as well as PRSs based on 660 asthma SNPs^46^, 8 rhinitis SNPs^47^, 425 eczema SNPs^48^, and 221 sensitization SNPs^39^.

##### Nasal and blood DNA methylation

the nasal and blood CpG sites were pre-selected based on EWAS summary statistics [FDR<0.05]^12,29,31^. After excluding CpGs which were not available in the discovery and replication cohorts, we included 112 nasal and 135 blood CpG sites in the analyses.

##### Sex and age

In total, 348 individuals had complete data on all the above features and were included in the model training.

All features can be founded in Table S13.

#### Machine learning method selection

We examined six supervised machine learning methods, including XGBoost^49^, Random Forest^50^, Support Vector Machine^51^, Naive Bayes^52^, Neural Network^53^, and Elastic Net^23^. Hyperparameter tuning was performed using Grid Search with ROC AUC as an evaluation metric, whereby each hyperparameter combination was evaluated using repeated cross-validation. Based on these results, we selected the method that provided accurate test performance, low overfitting, and good interpretability. The selected model was used for downstream analysis.

#### Model training

Models were estimated within a cross-validation framework, and 10-times repeated 10-fold cross-validation was adopted. This allowed for better model assessment, as it canceled out the randomness in fold splits and gave insight into performance variation over various runs^54^. To counteract the imbalance of the data (approx. 20% cases and 80% controls), different sampling techniques were assessed, including SMOTE^55^, downsampling and upsampling. The final model included upsampling in the training data, such that both classes had the same frequency by adding additional samples to the minority class with replacement. Test performance was defined as the average over the evaluation metric for the repeated cross-validation predictions, whereas model stability was assessed by inspecting the variation in performance of overall model runs. Besides the ROC curve, the Precision-Recall Curve (PRC) was used as an additional means for evaluating the model, to avoid the potentially misleading interpretation of ROC when working with imbalanced data^25^.

#### Predictive contribution of each data layer

The predictive contribution of each data layer is defined as the uplift in prediction performance after adding all its respective features to the model. We adhered to an iterative procedure, where layers were added sequentially. As the order of inclusion will affect the contribution found (because information can already be embedded in other data layers), we applied a sensitivity analysis on the order of the sequential addition.

### Generation of a parsimonious model

A more parsimonious model with fewer variables can lead to less overfitting and better prediction power. Firstly, the feature importance of all repeated cross-validation (cv) model runs was retrieved and the rank product statistic was used to aggregate this into a single feature with importance ranking based on all model runs^56^. Subsequently, we created a model with only the feature with the highest rank product statistic and its test performance was evaluated using 10-times repeated 10-fold cross-validation. The next most important feature was added iteratively to the model until no substantial further improvement in test performance was observed. Lastly, using the ‘corrected repeated k-fold cv test’^24^, the performance difference between incrementally more complex models was tested for significance. The final model was selected when the addition of more variables led to no significant improvement in model performance.

The 3-CpG model was also compared to the previously published 30-CpG nasal sites model that performed well in predicting atopy in the EVA-PR cohort^14^. We employed an Elastic Net model, including their prediction panel of 30 CpG sites, and made estimates with equal model specifications in terms of tuning and validation. Comparison was performed using the two-sided ‘corrected repeated k-fold cv test’.^24^

### External replication

After validating the reduced-feature model on the PIAMA cohort, we used the Epigenetic Variation and Childhood Asthma study in Puerto Ricans study (EVA-PR, including subjects aged 9 to 20 years)^14^ for replication, as well as two cohorts of younger children (mean age 6 years) COPSAC2010^57,58^ and MAKI^59^. Details of these cohorts are shown in the supplementary materials.

### Sensitivity analysis

To further assess the predictive power of the genetic factors, different significance thresholds for the selection of SNPs in the PRS construction were evaluated. This analysis was performed in a larger dataset of 675 individuals from the PIAMA cohort for whom both genotype and allergy phenotype data were available (see supplementary materials). Firstly, four different PRSs were calculated using SNPs selected by different p-value thresholds in the allergy GWAS^8^ (P < 1×10^−5^, 1×10^−6^, 1×10^−7^ and 5×10^−8^). Next, the prediction performance of each PRS was evaluated. Model performance was also examined for different subgroups of individuals stratified by IgE sensitization, for an allergy definition that is independent of sensitization and for sensitization itself as a target variable. We stratified the samples by IgE sensitization and symptom. Significant differences between each group-pair were assessed using a two-sided Student’s t-test.

### Biological interpretation of three CpG sites

To understand the function of the three CpG sites, we first annotated the CpG sites by position and then correlated the DNA methylation level to gene expression level by eQTM analysis. Briefly, 244 subjects had matched nasal DNA methylation and nasal gene expression data that were used for the eQTM analysis. We performed linear regression analysis to assess the association of the three CpG sites with all genes available, and the model was: gene expression level ∼ DNA methylation levels + age + sex + batch + study center. We controlled the FDR at 0.05 after multiple testing at each CpG level. We then performed WGCNA^26^ on the genes identified by eQTM analysis. Gene modules identified from WGCNA were used for KEGG pathway enrichment analysis using R package *topGO* (https://bioconductor.org/packages/release/bioc/html/topGO.html).

Single-cell RNA-seq data of nasal brushing samples were available from 4 subjects with asthma and 5 healthy controls, and were measured by 10x genomics. The detailed sample collection, alignment, QC, clustering and annotation steps have been reported previously^29^. In brief, cells with high percent.mito (>25%) and low gene counts (<500) were removed at each sample level. Ambient RNA was corrected using FastCAR (https://github.com/LungCellAtlas/FastCAR), and Scrublet^60^ was used for identifying doublets. Downstream analyses including normalization, scaling, clustering of cells and identifying cell marker genes were performed using the R package Seurat version 4.0 ^61^ (https://satijalab.org/seurat/). Harmony^62^ was used to integrate data from different donors.

In total, 10 cell type clusters (basal, goblet 1, goblet 2, squamous, cycling, ciliated, deuterosomal, ionocytes, myeloid and T cells) were identified. Re-clustering of the immune cell subset (myeloid and T cells) revealed the presence of two T cell clusters (CTL, and T cell cluster 2), three myeloid cell clusters (myeloid 1, myeloid 2 and macrophages), B cells, plasma cells and mast cells. To identify the sub-cluster of T cells and myeloid cells, we further re-clustered the two T cell clusters and the three myeloid cell clusters respectively. Three T cell clusters (CD8/ CTL effectors, CD4/ Treg and NK cells) and five myeloid cell clusters (DC-1, DC-2, macrophages, pDCs, classical monocytes) were identified. We then checked the expression pattern of eQTM genes from Module 1 and Module 2 (identified by WGCNA) in the identified cell types and immune cell subsets, by plotting scaled average expression of the genes in the different cell types.

Methylation quantitative trait loci (MeQTLs) analysis was performed using a linear model with R package *MatrixEQTL*^63^. In total, 433 subjects had matched genotype and DNA methylation data that were included in the MeQTL analysis. Mediation analysis was performed with R package *Mediation*^64^. Details of the methods can be found in the online supplementary materials.

## Data Availability

All data produced in the present study will be deposited in the European Genome-phenome Archive (EGA) upon publication.

## Data and code availability

Nasal and blood DNA methylation data from the discovery cohort (PIAMA) have been deposited in the European Genome-phenome Archive (EGA), which is hosted by the European Bioinformatics Institute (EMBL-EBI) and the Centre for Genomic Regulation (CRG), under accession number EGAS00001005189).

