## supplementary documents for "Nasal DNA methylation at three CpG sites predicts childhood allergic disease"

Merlijn van Breugel et al.

### Study and population description of discovery cohort

The discovery analysis was performed in the PIAMA birth cohort (Prevention and Incidence of Asthma and Mite Allergy). Details of the cohort have been published previously[1]. In brief, the national study started in 1996 with 3,963 newborns in the Netherlands. Questionnaire-based follow-up of the children took place at 3 months of age, annually from 1 to 8 years of age, and at 11, 14, and 16 years of age, with clinical investigations at ages 4, 8, 12 and 16 years. The Medical Ethical Committees of the participating institutes approved the study (Utrecht and Groningen METC (*Medisch Ethische Toetsings Commissie*) protocol number 12-019/K), and the parents and legal guardians of all participants, and later the participants themselves, gave written informed consent.

In this study, a case with allergic disease is a child with at least one of three allergic diseases (asthma/rhinitis/eczema) and who is specifically IgE positive (>0.35 lU/mL) to any allergen (house dust mite, cat, dactylis (grass) or birch). Asthma was defined as the presence of at least 2 of the following 3 criteria: 1) Doctor diagnosed asthma ever; 2) Wheeze in the last 12 months; and 3) Prescription of asthma medication in the last 12 months. Rhinitis was defined as the presence of sneezing or runny/blocked nose without having a cold in the last 12 months and nose symptoms accompanied by itchy, watering eyes. Eczema was defined as a positive answer to the question: has your child ever had an itchy rash which was coming and going in the last 12 months? If Yes: has this itchy rash affected any of the following places: the folds of the elbows, behind the knees, in front of the ankles, or around the neck, ears or eyes? Serum specific IgE to house dust mite, cat, dactylis (grass) and birch was measured and classified as positive if ≥ 0.35 IU/ml. Perinatal and environmental factors –including breastfeeding (yes/no), pets during pregnancy (yes/no), maternal smoking (yes/no), elder siblings at home (yes/no), and low birth weight (yes/no) – were assessed by questionnaires that were taken in the first year of life and regularly during follow-up.[1]

The initial dataset included 3,963 participants. The current study used the data of 16-year olds, and included validated questionnaires based on the harmonized MeDALL questionnaire[2], lung function measurements, blood testing for IgE sensitization, and DNA isolation of whole blood cells and nasal brushes. The medical examination at age 16 was restricted to children from the middle and northern regions of the Netherlands (Groningen and Utrecht) due to limited funding. We completed the examinations of in total 802 children at age 16 years. During the examination, blood and nasal samples were collected and used to extract DNA and RNA. In the 802 participants, 348 individuals had complete data of their clinical phenotype at 16 years old, genotype data, blood and nasal DNA methylation data, and environmental exposure data – these were all included in the discovery analysis of model generation (Table S9). In the sensitivity analysis of polygenic risk score (PRS), given the possibly limited prediction performance of genetics because of the small sample size, we tried to perform sensitivity analysis using a larger dataset from PIAMA that had both genotype and phenotype data at 16 years old (N = 675), regardless of the availability of other data layers. In the eQTM analysis, we included 244 participants that had both nasal DNA methylation and nasal RNA-seq data available. In the MeQTL analysis, we included 422 participants with both genotype and nasal DNA methylation data.

**Data measurement and quality control**

#### *Genotype data*

Genome-wide genotyping was performed in four phases. Quality control (QC) for each phase was performed and then the data were merged together. The first phase was performed within the framework of the GABRIEL Consortium, using an Illumina Human610 quad array. Genotypes were available from 363 children after QC. A second group of children were genotyped with an Illumina HumanOmniExpress array, with 272 individuals available after QC. A third group of children was genotyped with the Illumina Human Omni Express Exome Array, with 1333 individuals available after QC. A final group of children was genotyped with the Illumina Infinium Global Screening Array, with 107 individuals available after QC. SNPs were harmonized by base pair position annotated to genome build 37. In total, 2075 individuals remained after QC, and data from the four platforms were merged together.

#### *DNA methylation data*

We collected samples of peripheral blood, and of nasal epithelial cells by brushing. Briefly, the right nostril of the subjects was examined and the inferior turbinate was located using a speculum and penlight. Brushing was performed with a Cytosoft brush CP-5B (Cyto-Pak) after local anesthesia with 1% lidocaine spray. The lateral area underneath the inferior turbinate was then brushed for 3 seconds and the brush was placed in a 2 ml screw-cap Eppendorf tube and put into a freezer at -80°C until further processing. In total 4 brushes (2 for DNA isolation and 2 for RNA isolation) were collected.

DNA from whole blood was extracted using QIAamp blood kit (Qiagen Benelux BV, Venlo, the Netherlands) and nasal epithelium samples was extracted using DNA investigator kit (Qiagen Benelux BV). DNA concentration was determined by Nanodrop measurement and Picogreen quantification. 500 ng of DNA was bisulfite-converted using the EZ 96-DNA methylation kit (Zymo Research, Irvine, CA, USA), following the manufacturer's standard protocol. After verifying the bisulfite conversion step using Sanger sequencing, DNA concentration was normalized and the samples were randomized to avoid batch effects. One standard DNA sample per chip was included in this step for QC purposes.

Blood and nasal DNA samples were hybridized to the Infinium HumanMethylation450 BeadChip array (Illumina, San Diego, CA, USA). DNA methylation data were pre-processed in R with the Bioconductor package Minfi[3], using the original IDAT files extracted from the HiScanSQ scanner. We implemented sample filtering to remove poor quality samples (call rate <99%). Furthermore, we used the 65 SNP probes to check for concordances between paired DNA brush and blood samples from the same individuals. Paired samples that showed a SNP signal with a Pearson correlation coefficient <0.9 were regarded as sample mix-ups and were excluded from the study. We also verified the methylation distribution of the X-chromosome to verify gender. During processing, the probes on sex chromosomes, the probes that mapped to multiple loci, 65 SNP-probes and the probes containing SNPs at the target CpGs with a MAF >5% were excluded[4]. We implemented "DASEN"[5] to perform signal correction and normalization. After QC, 640 blood samples, 478 nasal samples, and 436,824 probes remained; after matching up with data from all the available layers, 348 samples were used for the analyses.

***RNA-sequencing data***

Total RNA was extracted using AllPrep DNA/RNA Mini kit (Qiagen Benelux BV). Samples were lysed in 600 µl RLT-plus buffer using an IKA Ultra Turrax T10 homogenizer, and RNA was purified according to the manufacturer's instructions. RNA samples were dissolved in 30 μl RNAse-free water. The concentrations and quality of RNA were checked using a Nanodrop ND-1000 and run on a Labchip GX (PerkinElmer Inc, Waltham, MA, USA).

Initial QC and RNA quantification of the samples was performed by capillary electrophoresis using the LabChip GX (PerkinElmer, Inc). Non-degraded RNA-samples with integrity scores RIN > 6 were selected for subsequent sequencing analysis. Sequence libraries were generated by Poly (A) enrichment using the TruSeq RNA Sample Prep Kit (Illumina) using the Sciclone NGS Liquid Handler (PerkinElmer). In case of contamination of adapter-duplexes, an extra purification of the libraries was performed with the automated agarose gel separation system, Labchip XT (PerkinElmer). Whenever sample preparation failed, either a new sample preparation was attempted or the sample was replaced by a spare nose brush sample from the same individual. The cDNA fragment libraries obtained were loaded in pools of multiple samples into an Illumina HiSeq2500 sequencer using default parameters for paired-end sequencing (2 × 100 bp). If sequencing of a library gave insufficient reads, a second sequencing run was performed to generate additional reads; our target was 15 million read-pairs per sample.

The trimmed fastQ files where aligned to build b37 of the human reference genome using HISAT (version 0.1.5) [6], allowing for 2 mismatches. Reads from separate runs of the same library were merged. Before gene quantification, SAMtools (version 1.2) was used to sort the aligned reads[7]. The gene level quantification was performed by HTSeq (version 0.6.1p1) using –mode=union --stranded=no and using Ensembl version 75 as the gene annotation database[8].

Quality control metrics were calculated for the raw sequencing data using the FastQC tool (version 0.11.3). Alignments of RNA of 333 subjects were obtained. QC metrics were calculated for the aligned reads using Picard-tools (version 1.130) *CollectRnaSeqMetrics, MarkDuplicates, CollectInsertSize-Metrics and SAMtools flagstat* (http://picard.sourceforge.net). We discarded five samples without replacement due to poor alignment metrics. In addition, we checked for concordance between sex-linked (XIST and Y-chromosomal genes) gene expression and reported sex. Two more samples were discarded for lacking concordance. This resulted in high quality RNA-seq data from 326 subjects.

Expressed features were excluded from analysis if fewer than half the samples had counts per million mapped reads (CPM) of at least 5/M, where M is the median library size in millions. This left 17,156 expressed features for analysis. Raw count data were transformed to log2CPM using *voom* and analyzed in the *limma* package[9].

**External replication**

We first replicated our model in a cohort of comparable age but different ethnicity (Epigenetic Variation and Childhood Asthma in Puerto Ricans (EVA-PR).

***EVA-PR***

The Epigenetic Variation and Childhood Asthma in Puerto Ricans (EVA-PR) is a case-control study of asthma in subjects aged 9-20 years; cohort recruitment, procedures, and methods have been described previously[10,11]. Briefly, participants with and without asthma were recruited from randomly selected households in San Juan (PR) from February 2014 through May 2017, using multistage probability sampling. The study was approved by the institutional review boards of the University of Puerto Rico (San Juan, PR) and the

University of Pittsburgh (Pittsburgh, PA, USA). Written parental consent and assent from participants <18 years old were obtained, and consent was obtained from participants ≥18 years old. The study protocol included questionnaires on respiratory health, measurement of serum allergen-specific IgE, and taking nasal samples for DNA extraction. Asthma was defined as physician-diagnosed asthma with at least one episode of wheezing in the previous year. Allergic rhinitis was defined as hay fever or naso-ocular symptoms (a runny or stuffy nose accompanied by sneezing and itching) apart from a cold or flu in the previous 12 months. Eczema was defined as a prolonged, itchy, scaly or weepy skin rash in the previous 12 months. Atopy was defined as ≥1 positive IgE (⩾0.35 IU/mL) to at least one of five common aeroallergens in Puerto Rico: house dust mite, cockroach, cat dander, dog dander, or mouse urinary protein. Allergic disease was defined as the presence of asthma or allergic rhinitis or eczema AND atopy. DNA was extracted from nasal specimens collected from the inferior turbinate. Whole-genome methylation assays were performed using HumanMethylation450 BeadChips (Illumina). After QC, 227,836 CpG probes remained. Methylation β-values were calculated as a percentage: $\beta=M/(M+U+\alpha)$, where M and U represent methylated and unmethylated signal intensities, respectively, and α is an arbitrary offset to stabilize β-values if fluorescent intensities were low. β-values were then transformed to M-values as $\log_2(\beta/(1-\beta))$, and M-values were used in all downstream analyses. Known batch effects (e.g. plates) were removed using an empirical Bayes framework, and *sva* was used to estimate latent factors (LFs) that capture unknown data heterogeneity. To account for population stratification, we also adjusted our models for principal components derived from genotype data (using Illumina HumanOmni2.5 BeadChips).

We also tried to expand the model to other ages especially to two younger cohorts of children aged around 6 years: Copenhagen prospective studies on asthma in childhood (COPSAC)[12] and the Dutch MAKI trial[13].

***COPSAC***

The $COPSAC_{2010}$ cohort is an ongoing, prospective mother-child cohort comprising 731 children born to unselected mothers from Zealand, Denmark, in 2009-2010 as described previously[12]. At week 24 of pregnancy, women were randomly assigned to receive n-3 long chain polyunsaturated fatty acids (n = 362) and placebo (n = 369). Baseline information as well as details of this randomized controlled trial have been published[14].

The study was conducted in accordance with the guiding principles of the Declaration of Helsinki and was approved by the Local Ethics Committee (COPSAC2010: H-B-2008-093), and the Danish Data Protection Agency ($COPSAC_{2010}$: 2015-41-3696). Both parents provided written informed consent before enrolment.

RNA was collected from inferior turbinate epithelial cell scrapings from 562 children. Samples were obtained using a rhino-probe nasal curette, stored in RNAprotect Cell Reagent (Qiagen, Germantown, MD, USA), and then cryopreserved at −80°C until nucleotide extractions could be performed[15]. After removing samples that did not pass QC or without genotype information (n = 59), 503 samples remained.

Out of 866,836 probes on the Illumina Infinium Methylation EPIC array, we removed 23,172 with a detection p-value of > 0.01 in 90% of the samples, 18,695 located on sex chromosomes, and 120,903 probes that overlapped a known common SNP (MAF > 0.05) or mapped to multiple positions as a result of cross-hybridization. 703,565 CpG sites passed QC and were used in downstream analyses. Normalization was performed using the SWAN algorithm from the R package *minfi* (version 1.26) and quantile normalization from the R package *lumi* (version 2.36). Principal component analysis (PCA) was used to identify the effect of confounding variables on DNA methylation: DNA concentration, slide and site of sampling significantly correlated with at least one of the first ten principal components and were included in the final model.

***MAKI***

The MAKI trial enrolled 429 otherwise healthy, late preterm infants between 2008 and 2010; they were born at 33 to 35 weeks gestation. Infants were randomly assigned to receive palivizumab (n = 214) or placebo (n = 215) during their first RSV season. Baseline characteristics at randomization are available at NEJM.org[13]. Details about the design, definitions, protocol of the primary study, follow-up study and clinical assessment have been previously described[13,16].

Nasal epithelial cells were collected by brushing around age 6. Briefly, the right nostril of the subjects was examined and the inferior turbinate was located using a speculum and penlight. The lateral area underneath the inferior turbinate was then brushed for 3 seconds with two brushes (Copan, 56380CS01 FLOQswabs) and these were placed in a 2 ml screw-cap Eppendorf tube and put into a freezer at – 80o C until further processing.

DNA was extracted from nasal brushes using the DNA investigator kit (Qiagen, Benelux BV, Venlo, the Netherlands). This was followed by precipitation-based purification and concentration using GlycoBlue (Ambion). 500 ng of DNA was bisulfite-converted using the EZ 96-DNA methylation kit (Zymo Research), following the manufacturer’s standard protocol. After verification of the bisulfite conversion step using Sanger Sequencing, DNA concentration was normalized and the samples were randomized to avoid batch effects. One standard DNA sample per chip was included in this step for quality control. Study personnel and technicians were blinded for the intervention.

In total, 296 nasal epithelium samples with sufficient DNA quality and quantity were hybridized to the Infinium HumanMethylationEPIC BeadChip array (Illumina, San Diego, CA, USA). DNA methylation data were pre-processed in R (version 3.3.2) with the Bioconductor package *Minfi*, using the original IDAT files extracted from the HiScanSQ scanner. We implemented sample filtering to remove 6 bad quality samples (call rate <99%) and 16 samples with gender mismatch. During processing, bad quality probes which failed more than 10% of the samples, the probes on sex chromosomes, the probes that mapped to multiple loci, and the probes containing SNPs at the target CpG sites with a MAF>5% in European populations were excluded. We subsequently implemented stratified quantile normalization. After quality control, 274 (63.8%) of the collected samples and 790,437 probes remained for further analyses.

***Nasal single cell RNA-seq cohort***

Nasal brush samples from 4 asthma patients and 5 healthy controls were collected from inferior turbinate by Cytosoft brush CP-5B (Cyto-Pak). The brushes were collected in a 50mL tube containing HBSS(Lonza) + 1%Pen/Strep. Cells were spinned down at 560xg for 5 min. Cell pellet was then resuspended in HBSS containing 1mg/ml Collagenase D and 0.1mg/ml DNase I (Roche) and placed at 37°C for 1 hour with gentle agitation. Cell suspension was pushed through a 70uM nylon cell strainer (Falcon) and spinned down at 560xg for 5 min. Next, cells were washed with PBS containing 1% BSA (Sigma Aldrich).

Single cell suspension was cleared of red blood cells using a Red Blood cell lysis buffer (eBioscience).

Cell suspension was counted manually using a haemocytometer and concentration was adjusted to a minimum of 300 cells/ul. Cells were loaded according to the standard protocol of Chromium single cell 3'kit. Following steps were performed according to Single Cell 3'Reagent Kits V2 User guide. We performed RNA-sequencing on Illumina Hiseq 4000 or NOVA-Seq 6000 aiming to achieve a mean coverage of 50k -100k reads/cell. 10X Genomics raw sequencing data was processed using CellRanger software and the 10X human genome GRCh38 1.2.0 release as the reference. Downstream analyses including clustering of cells and identifying cluster marker genes were performed using the R software package Seurat version 4 (https://github.com/satijalab/seurat).

### Supplementary figures S1-S7

**Nasal DNA methylation at three CpG sites predicts childhood allergic disease**

M. van Breugel et al.

**Figure S1. Sensitivity analysis of PRS.** (a) Differential analysis of polygenic risk scores (PRS) using different p-value thresholds. (b) PRS are calculated using matched summary statistics of disease-specific GWAS (allergy, asthma, eczema, rhinitis and IgE sensitization), using a p-value threshold of 1e$^{-7}$ for SNP selection. No significant differences in the distribution between case and control groups was observed for the phenotypes (Student's t-test, ns: p-value > 0.05).

(a)

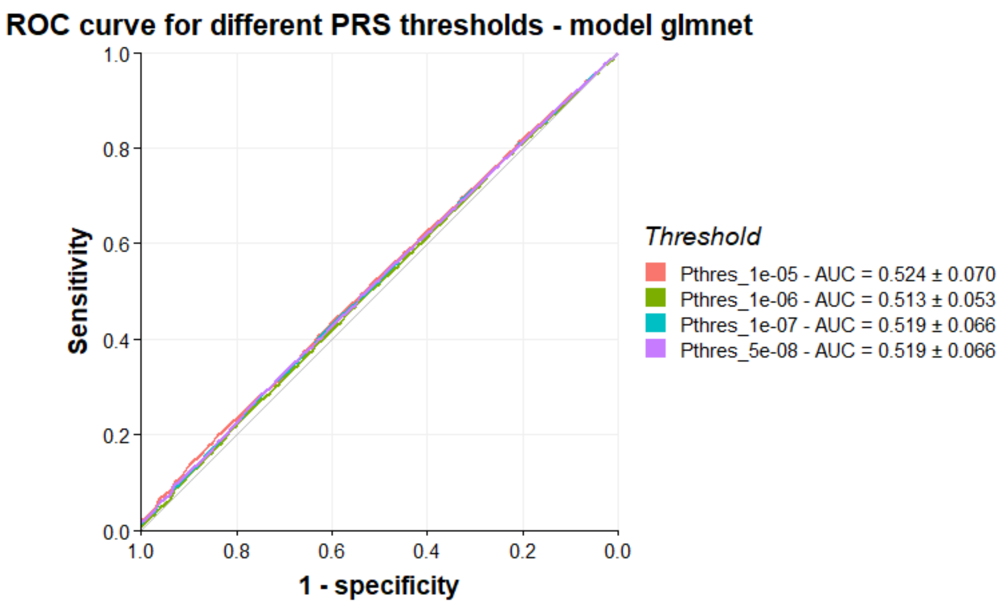


(b)

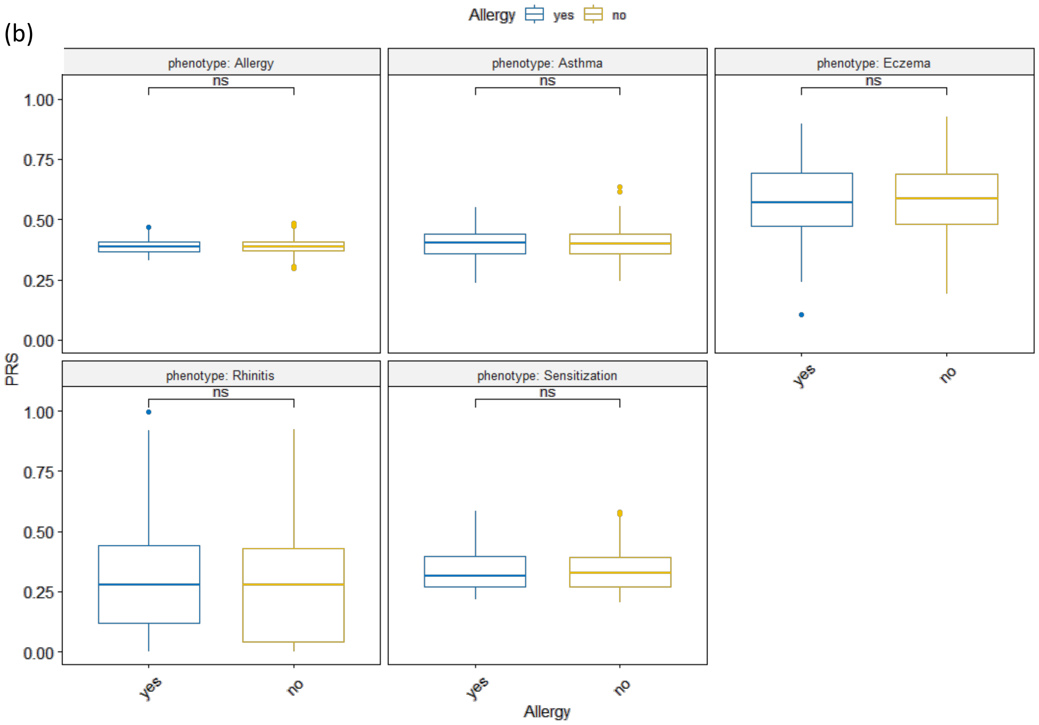

**Figure S2. Precision-recall curves (PRC) for the 3-CpG sites model in the discovery cohort (a) and replication cohorts (b EVA-PR; c COPSAC; d MAKI).**

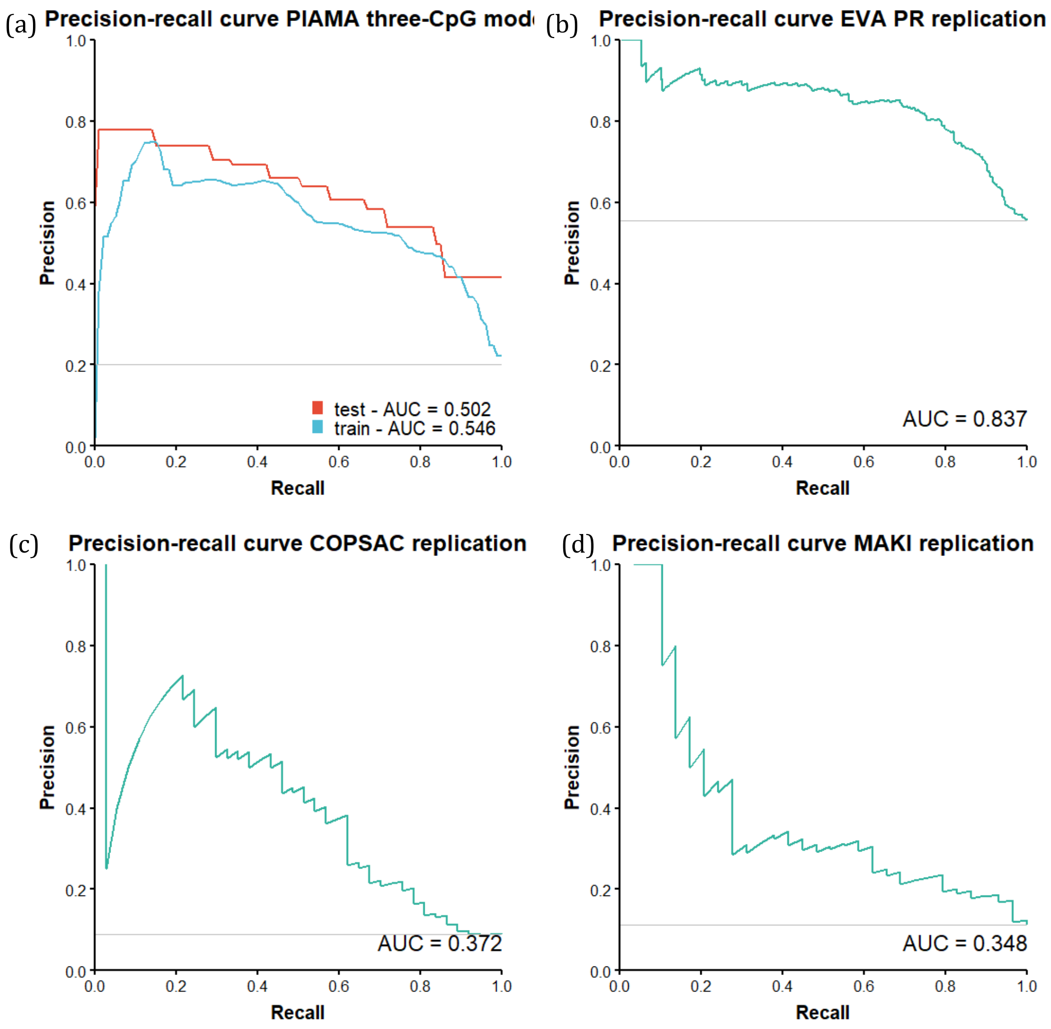

**Figure S3. Performance curves of Forno et al.'s (2019) prediction panel**, using their 30 CpG sites. These sites are used as model variables for an Elastic Net, which is estimated in the standard 10-times repeated 10-fold cross-validation framework. Their model has been tuned using the same parameters as for the discovery cohort. (a) ROC curve, (b) PRC curve.

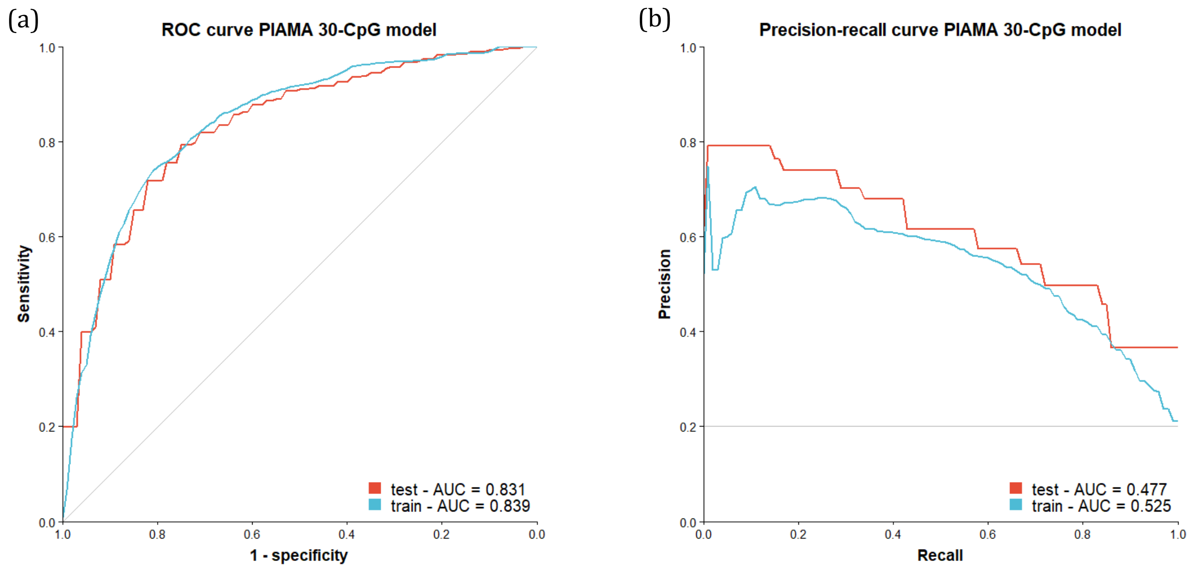


Ref: Forno, E. *et al.* DNA methylation in nasal epithelium, atopy, and atopic asthma in children: a genome-wide study. *Lancet Respir. Med.* 7, 336–346 (2019).

**Figure S4. Heatmap of eQTM results.** Each column is gene identified by eQTM analysis; on the y-axis are the three CpG sites in the model. Red indicates a positive correlation between a CpG and a gene; blue indicates a negative correlation. The color bars on the top indicate the significance level (yes, FDR<0.05; no, FDR>=0.05) of each gene associated with each CpG site.

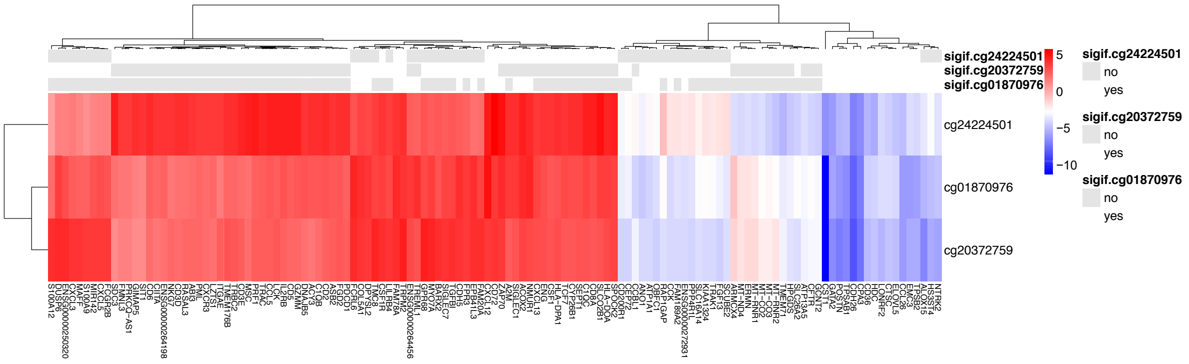

**Figure S5. KEGG pathway enrichment of two gene modules identified by WGCNA.** We identified two gene modules from eQTM genes by Weighted Gene Co-expression Network Analysis (WGCNA). The bubble plot shows the results of KEGG pathway enrichment analysis for module 1 (a) and for module 2 (b).

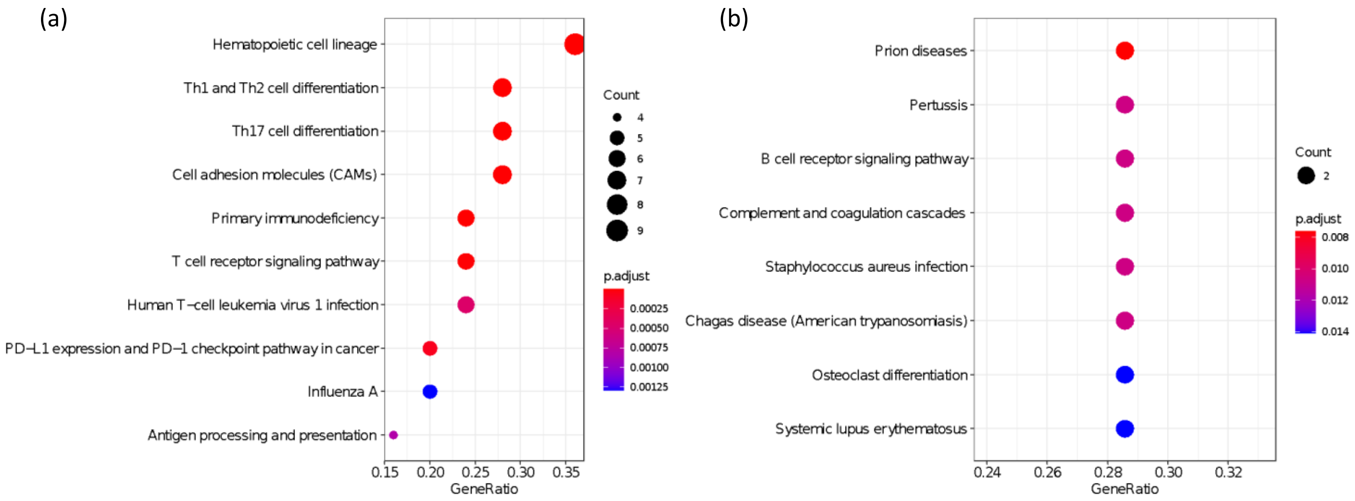

**Figure S6. DNA methylation levels of three CpG sites in nasal brushes and blood in the PIAMA birth cohort**

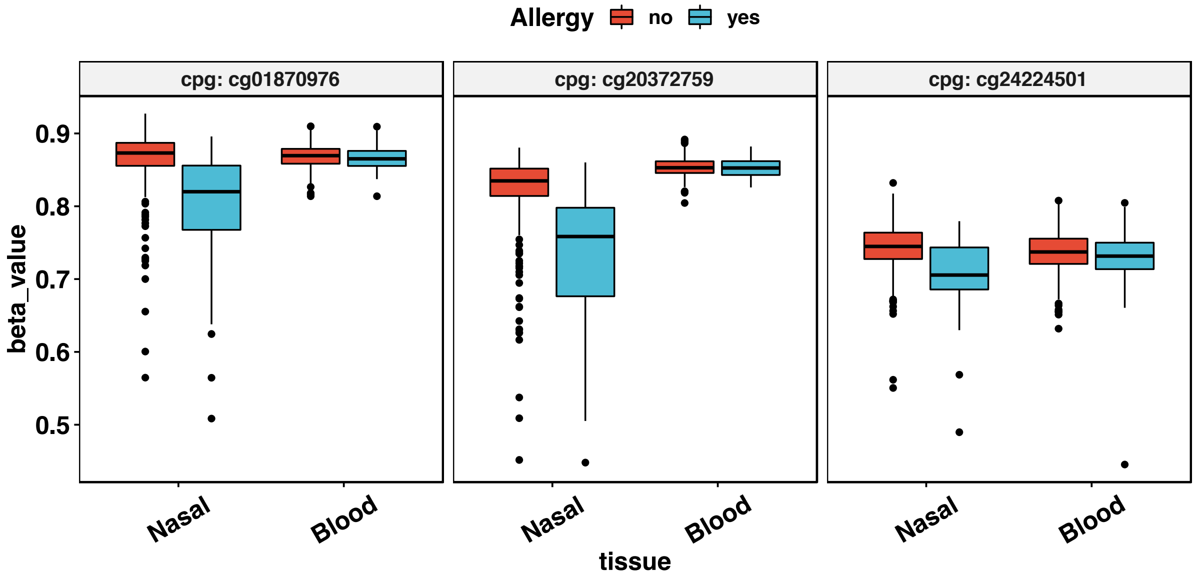

**Figure S7. Venn diagram showing samples with both allergy symptom and IgE sensitization (the allergy phenotype used in this study) (a), and samples with allergy symptom but without IgE sensitization (b).**

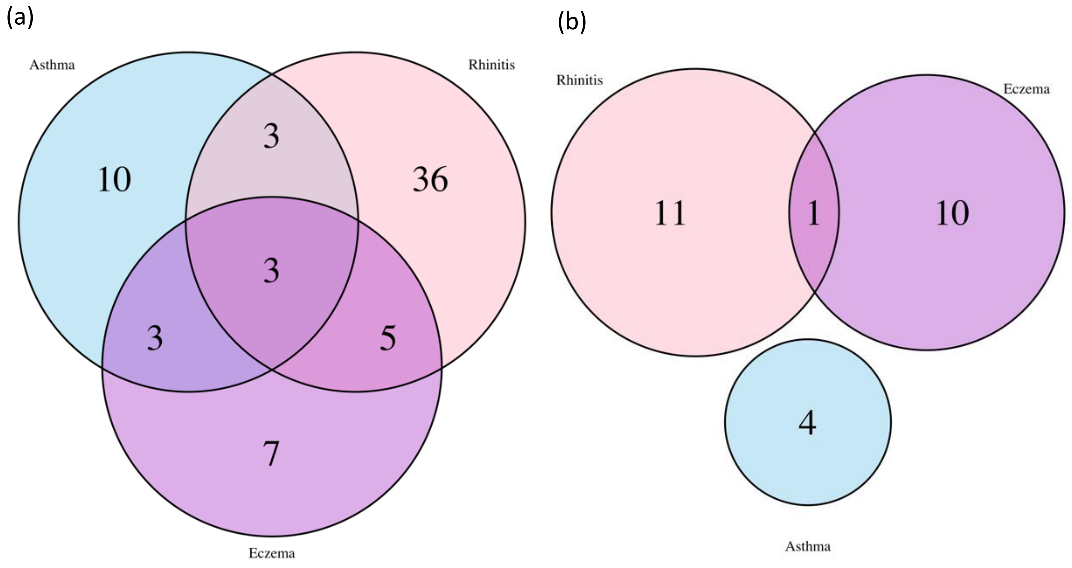

Table S1. Comparison of prediction performance for different machine learning techniques

| | Training | | | | Testing | | | |
|---|---|---|---|---|---|---|---|---|
| Model | ROC AUC mean | SD | PRC AUC mean | SD | ROC AUC mean | SD | PRC AUC mean | SD |
| Elastic Net | 0,874 | 0,009 | 0,560 | 0,020 | 0,835 | 0,079 | 0,470 | 0,126 |
| Naive Bayes | 0,894 | 0,179 | 0,693 | 0,266 | 0,720 | 0,132 | 0,345 | 0,143 |
| Neural Net (1-layer) | 0,931 | 0,037 | 0,733 | 0,100 | 0,730 | 0,099 | 0,383 | 0,115 |
| Random Forest | 1,000 | 0,000 | 0,983 | 0,000 | 0,838 | 0,076 | 0,464 | 0,130 |
| Support Vector Machine (Radial kernel) | 0,904 | 0,013 | 0,636 | 0,026 | 0,799 | 0,090 | 0,443 | 0,128 |
| XGBoost | 0,988 | 0,013 | 0,943 | 0,041 | 0,826 | 0,077 | 0,464 | 0,127 |

ROC AUC: receiver operating characteristic, area under curve

PRC AUC: precision-recall curve, area under curve

Table S2. Summary of ranking of features from different data layers

| | | Feature ranking | | | |
|---|---|---|---|---|---|
| Daya layer | Full feature model (No.) | No. in top 150 (% of top X) | No. in top 100 (% of top X) | No. in top 50 (% of top X) | No. in top 10 (% of top X) |
| Host | 2 | 1 (0.7%) | 0 (0%) | 0 (0%) | 0 (0%) |
| Environment | 4 | 2 (1.3%) | 0 (0%) | 0 (0%) | 0 (0%) |
| Perinatal | 2 | 0 (0%) | 0 (0%) | 0 (0%) | 0 (0%) |
| Genetics | 106 | 3 (2%) | 1 (1%) | 0 (0%) | 0 (0%) |
| Blood DNA methylation | 219 | 76 (50.7%) | 43 (43%) | 7 (14%) | 0 (0%) |
| Nasal DNA methylation | 134 | 68 (45.3%) | 56 (56%) | 43 (86%) | 10 (100%) |

Host: age and gender

Environment: cats or dogs during pregnancy, maternal smoking (mother smoking in pregnancy but stopped between 12 and 16 weeks and mother smoking in pregnancy at 16 weeks), older siblings at home

Perinatal: low birth weight and breastfeeding

Genetics: allergy SNP dosages and polygenic risk scores (PRS) for the combined allergy phenotype, as well as for asthma, rhinitis, eczema and IgE-sensitization

Blood methylation: 219 CpG sites from blood cells that were previously associated with allergy

Nasal methylation: 134 CpG sites from nasal cells that were previously associated with allergy

Table S3. Rank products of top-10 ranked features

| Rank | Feature | Rank product | AUROC |
|---|---|---|---|
| 1 | methylNasal - cg20372759 | 2963,495 | 0,816 |
| 2 | methylNasal - cg01870976 | 1717,827 | 0,824 |
| 3 | methylNasal - cg24224501 | 1541,129 | 0,856 |
| 4 | methylNasal - cg22862094 | 1454,589 | 0,854 |
| 5 | methylNasal - cg16027132 | 1441,167 | 0,848 |
| 6 | methylNasal - cg20790648 | 1218,264 | 0,855 |
| 7 | methylNasal - cg24707200 | 1070,770 | 0,852 |
| 8 | methylNasal - cg15006973 | 921,767 | 0,854 |
| 9 | methylNasal - cg08844313 | 917,173 | 0,855 |
| 10 | methylNasal - cg22689016 | 714,923 | 0,850 |

Table S4. Annotation of three CpG sites in our parsimonious model

| CpG | CHR | BP | GREAT annotation |
|---|---|---|---|
| cg20372759 | 12 | 58162287 | METTL21B(-4095),CYP27B1(-1254) |
| cg01870976 | 15 | 101887154 | SNRPA1(-51699),PCSK6(+142718) |
| cg24224501 | 7 | 102566723 | ARMC10(-148604),LRRC17(+13286) |

Table S5. Three-CpG model specification

| Feature name | Coefficient |
|---|---|
| (Intercept) | -0,0025882 |
| methylNasal - cg01870976 | 0,11658298 |
| methylNasal - cg20372759 | 0,13323378 |
| methylNasal - cg24224501 | 0,09059412 |

Coefficients for three-CpG model of Elastic Net model, resulting after hyperparameter optimization on PIAMA cohort using the *caret* package in R (alpha=0.1, lambda=0.9)

Table S6. Performance metrics for all four cohorts

| | PIAMA | EVA_PR | COPSAC | MAKI |
|---|---|---|---|---|
| Accuracy | 0,815 | 0,771 | 0,921 | 0,892 |
| Sensitivity | 0,638 | 0,787 | 0,270 | 0,034 |
| Specificity | 0,857 | 0,752 | 0,984 | 1,000 |
| Pos Pred Value | 0,533 | 0,798 | 0,625 | 1,000 |
| Neg Pred Value | 0,910 | 0,739 | 0,933 | 0,891 |
| Precision | 0,533 | 0,798 | 0,625 | 1,000 |
| Recall | 0,638 | 0,787 | 0,270 | 0,034 |
| F1 | 0,569 | 0,792 | 0,377 | 0,067 |

Table S7. Results of eQTM analysis for three CpG sites in nasal brushes

| CpG | gene | hgnc_symbol | coef | t | P.Value | fdr |
|---|---|---|---|---|---|---|
| cg20372759 | ENSG00000170373 | CST1 | -3,211 | -11,282 | 5,70E-24 | 2,93E-19 |
| cg01870976 | ENSG00000170373 | CST1 | -3,421 | -11,150 | 1,52E-23 | 3,91E-19 |
| cg20372759 | ENSG00000124215 | CDH26 | -0,691 | -8,507 | 1,92E-15 | 3,30E-11 |
| cg01870976 | ENSG00000124215 | CDH26 | -0,707 | -8,011 | 4,92E-14 | 6,33E-10 |
| cg20372759 | ENSG00000163751 | CPA3 | -2,537 | -7,390 | 2,42E-12 | 2,49E-08 |
| cg01870976 | ENSG00000163751 | CPA3 | -2,631 | -7,083 | 1,55E-11 | 1,18E-07 |
| cg20372759 | ENSG00000133110 | POSTN | -1,700 | -7,077 | 1,61E-11 | 1,18E-07 |
| cg24224501 | ENSG00000124215 | CDH26 | -1,226 | -6,890 | 4,86E-11 | 3,13E-07 |
| cg20372759 | ENSG00000172236 | TPSAB1 | -2,137 | -6,844 | 6,35E-11 | 3,63E-07 |
| cg20372759 | ENSG00000161905 | ALOX15 | -0,483 | -6,702 | 1,45E-10 | 7,46E-07 |
| cg24224501 | ENSG00000163751 | CPA3 | -4,888 | -6,646 | 2,00E-10 | 9,34E-07 |
| cg01870976 | ENSG00000172236 | TPSAB1 | -2,211 | -6,550 | 3,45E-10 | 1,48E-06 |
| cg20372759 | ENSG00000179348 | GATA2 | -1,670 | -6,532 | 3,83E-10 | 1,52E-06 |
| cg24224501 | ENSG00000170373 | CST1 | -4,357 | -6,361 | 1,00E-09 | 3,69E-06 |
| cg01870976 | ENSG00000133110 | POSTN | -1,649 | -6,274 | 1,62E-09 | 5,57E-06 |
| cg24224501 | ENSG00000179348 | GATA2 | -3,336 | -6,150 | 3,20E-09 | 1,03E-05 |
| cg20372759 | ENSG00000006606 | CCL26 | -1,131 | -6,115 | 3,89E-09 | 1,18E-05 |
| cg01870976 | ENSG00000006606 | CCL26 | -1,210 | -6,090 | 4,44E-09 | 1,27E-05 |
| cg01870976 | ENSG00000125037 | EMC3 | -0,184 | -6,008 | 6,92E-09 | 1,87E-05 |
| cg01870976 | ENSG00000197253 | TPSB2 | -2,028 | -5,931 | 1,04E-08 | 2,68E-05 |
| cg01870976 | ENSG00000179348 | GATA2 | -1,612 | -5,777 | 2,34E-08 | 5,73E-05 |
| cg24224501 | ENSG00000137101 | CD72 | 0,995 | 5,758 | 2,58E-08 | 6,04E-05 |
| cg20372759 | ENSG00000148053 | NTRK2 | -0,956 | -5,727 | 3,04E-08 | 6,79E-05 |
| cg20372759 | ENSG00000182601 | HS3ST4 | -1,156 | -5,649 | 4,55E-08 | 9,36E-05 |
| cg24224501 | ENSG00000140287 | HDC | -1,813 | -5,655 | 4,41E-08 | 9,36E-05 |
| cg20372759 | ENSG00000125037 | EMC3 | -0,160 | -5,546 | 7,67E-08 | 1,46E-04 |
| cg20372759 | ENSG00000197253 | TPSB2 | -1,782 | -5,551 | 7,48E-08 | 1,46E-04 |
| cg24224501 | ENSG00000133110 | POSTN | -2,790 | -5,308 | 2,52E-07 | 4,63E-04 |
| cg01870976 | ENSG00000161905 | ALOX15 | -0,421 | -5,276 | 2,95E-07 | 5,24E-04 |
| cg24224501 | ENSG00000135218 | CD36 | -1,034 | -5,253 | 3,29E-07 | 5,65E-04 |
| cg20372759 | ENSG00000170500 | LONRF2 | -0,597 | -5,175 | 4,80E-07 | 7,97E-04 |
| cg01870976 | ENSG00000107562 | CXCL12 | 1,138 | 5,157 | 5,24E-07 | 8,37E-04 |
| cg24224501 | ENSG00000165071 | TMEM71 | -0,977 | -5,152 | 5,36E-07 | 8,37E-04 |
| cg24224501 | ENSG00000115085 | ZAP70 | 1,051 | 5,134 | 5,85E-07 | 8,85E-04 |
| cg20372759 | ENSG00000135218 | CD36 | -0,480 | -5,113 | 6,48E-07 | 9,42E-04 |
| cg24224501 | ENSG00000137491 | SLCO2B1 | 0,955 | 5,109 | 6,59E-07 | 9,42E-04 |
| cg24224501 | ENSG00000172236 | TPSAB1 | -3,464 | -5,078 | 7,63E-07 | 1,06E-03 |
| cg01870976 | ENSG00000182601 | HS3ST4 | -1,082 | -4,849 | 2,22E-06 | 3,01E-03 |
| cg24224501 | ENSG00000107562 | CXCL12 | 2,105 | 4,838 | 2,35E-06 | 3,10E-03 |
| cg20372759 | ENSG00000012660 | ELOVL5 | -0,288 | -4,813 | 2,63E-06 | 3,31E-03 |
| cg24224501 | ENSG00000006606 | CCL26 | -1,925 | -4,818 | 2,57E-06 | 3,31E-03 |
| cg20372759 | ENSG00000109861 | CTSC | -0,246 | -4,796 | 2,84E-06 | 3,48E-03 |
| cg24224501 | ENSG00000180644 | PRF1 | 1,406 | 4,788 | 2,94E-06 | 3,52E-03 |
| cg24224501 | ENSG00000107742 | SPOCK2 | 0,838 | 4,733 | 3,78E-06 | 4,42E-03 |
| cg24224501 | ENSG00000178860 | MSC | 1,442 | 4,703 | 4,32E-06 | 4,94E-03 |
| cg24224501 | ENSG00000182866 | LCK | 0,938 | 4,692 | 4,54E-06 | 5,08E-03 |
| cg24224501 | ENSG00000159189 | C1QC | 0,980 | 4,676 | 4,87E-06 | 5,22E-03 |
| cg24224501 | ENSG00000197540 | GZMM | 1,528 | 4,680 | 4,80E-06 | 5,22E-03 |
| cg01870976 | ENSG00000148053 | NTRK2 | -0,849 | -4,637 | 5,81E-06 | 5,34E-03 |
| cg20372759 | ENSG00000119714 | GPR68 | 0,533 | 4,640 | 5,73E-06 | 5,34E-03 |

| | | | | | | |
|---|---|---|---|---|---|---|
| cg20372759 | ENSG00000140287 | HDC | -0,721 | -4,638 | 5,77E-06 | 5,34E-03 |
| cg24224501 | ENSG00000012660 | ELOVL5 | -0,585 | -4,638 | 5,78E-06 | 5,34E-03 |
| cg24224501 | ENSG00000100385 | IL2RB | 1,085 | 4,652 | 5,44E-06 | 5,34E-03 |
| cg24224501 | ENSG00000110448 | CD5 | 0,897 | 4,642 | 5,67E-06 | 5,34E-03 |
| cg24224501 | ENSG00000153563 | CD8A | 1,194 | 4,662 | 5,20E-06 | 5,34E-03 |
| cg24224501 | ENSG00000210082 | MT-RNR2 | -0,681 | -4,637 | 5,81E-06 | 5,34E-03 |
| cg24224501 | ENSG00000162512 | SDC3 | 0,880 | 4,620 | 6,25E-06 | 5,65E-03 |
| cg24224501 | ENSG00000161570 | CCL5 | 1,308 | 4,613 | 6,45E-06 | 5,73E-03 |
| cg24224501 | ENSG00000168306 | ACOX2 | 0,596 | 4,601 | 6,80E-06 | 5,93E-03 |
| cg20372759 | ENSG00000142185 | TRPM2 | 0,459 | 4,588 | 7,21E-06 | 6,18E-03 |
| cg01870976 | ENSG00000131620 | ANO1 | -0,548 | -4,541 | 8,84E-06 | 7,46E-03 |
| cg24224501 | ENSG00000163106 | HPGDS | -2,069 | -4,536 | 9,05E-06 | 7,52E-03 |
| cg01870976 | ENSG00000181036 | FCRL6 | 0,765 | 4,501 | 1,05E-05 | 8,32E-03 |
| cg20372759 | ENSG00000137474 | MYO7A | 0,457 | 4,501 | 1,05E-05 | 8,32E-03 |
| cg24224501 | ENSG00000198851 | CD3E | 0,961 | 4,502 | 1,05E-05 | 8,32E-03 |
| cg24224501 | ENSG00000229164 | TRAC | 0,898 | 4,498 | 1,07E-05 | 8,32E-03 |
| cg01870976 | ENSG00000171596 | NMUR1 | 0,861 | 4,494 | 1,09E-05 | 8,34E-03 |
| cg24224501 | ENSG00000204252 | HLA-DOA | 0,986 | 4,490 | 1,10E-05 | 8,36E-03 |
| cg24224501 | ENSG00000198938 | MT-CO3 | -0,642 | -4,423 | 1,47E-05 | 1,10E-02 |
| cg20372759 | ENSG00000167895 | TMC8 | 0,302 | 4,415 | 1,53E-05 | 1,12E-02 |
| cg20372759 | ENSG00000129682 | FGF13 | -0,410 | -4,406 | 1,59E-05 | 1,15E-02 |
| cg24224501 | ENSG00000013725 | CD6 | 0,829 | 4,393 | 1,67E-05 | 1,20E-02 |
| cg24224501 | ENSG00000111846 | GCNT2 | -0,516 | -4,388 | 1,71E-05 | 1,21E-02 |
| cg20372759 | ENSG00000139318 | DUSP6 | 0,357 | 4,384 | 1,74E-05 | 1,21E-02 |
| cg01870976 | ENSG00000126882 | FAM78A | 0,412 | 4,381 | 1,77E-05 | 1,21E-02 |
| cg24224501 | ENSG00000109861 | CTSC | -0,474 | -4,366 | 1,88E-05 | 1,27E-02 |
| cg01870976 | ENSG00000135218 | CD36 | -0,445 | -4,353 | 1,99E-05 | 1,33E-02 |
| cg24224501 | ENSG00000155850 | SLC26A2 | -0,853 | -4,328 | 2,21E-05 | 1,46E-02 |
| cg20372759 | ENSG00000107960 | OBFC1 | -0,158 | -4,307 | 2,41E-05 | 1,55E-02 |
| cg24224501 | ENSG00000171596 | NMUR1 | 1,623 | 4,307 | 2,41E-05 | 1,55E-02 |
| cg01870976 | ENSG00000168306 | ACOX2 | 0,285 | 4,297 | 2,51E-05 | 1,60E-02 |
| cg24224501 | ENSG00000001626 | CFTR | -0,856 | -4,288 | 2,61E-05 | 1,64E-02 |
| cg20372759 | ENSG00000250320 | | 0,773 | 4,273 | 2,79E-05 | 1,73E-02 |
| cg24224501 | ENSG00000116824 | CD2 | 0,908 | 4,261 | 2,92E-05 | 1,79E-02 |
| cg01870976 | ENSG00000163399 | ATP1A1 | -0,156 | -4,247 | 3,10E-05 | 1,88E-02 |
| cg01870976 | ENSG00000171097 | CCBL1 | -0,387 | -4,233 | 3,28E-05 | 1,96E-02 |
| cg01870976 | ENSG00000115085 | ZAP70 | 0,448 | 4,225 | 3,39E-05 | 1,97E-02 |
| cg01870976 | ENSG00000140287 | HDC | -0,710 | -4,223 | 3,42E-05 | 1,97E-02 |
| cg01870976 | ENSG00000161911 | TREML1 | 0,815 | 4,216 | 3,52E-05 | 1,97E-02 |
| cg20372759 | ENSG00000043039 | BARX2 | 0,495 | 4,216 | 3,53E-05 | 1,97E-02 |
| cg20372759 | ENSG00000181036 | FCRL6 | 0,672 | 4,225 | 3,40E-05 | 1,97E-02 |
| cg24224501 | ENSG00000146192 | FGD2 | 0,729 | 4,219 | 3,48E-05 | 1,97E-02 |
| cg20372759 | ENSG00000082397 | EPB41L3 | 0,504 | 4,202 | 3,74E-05 | 2,07E-02 |
| cg24224501 | ENSG00000106565 | TMEM176B | 0,993 | 4,189 | 3,93E-05 | 2,13E-02 |
| cg24224501 | ENSG00000132744 | ACY3 | 1,255 | 4,192 | 3,89E-05 | 2,13E-02 |
| cg24224501 | ENSG00000188389 | PDCD1 | 1,322 | 4,187 | 3,98E-05 | 2,13E-02 |
| cg20372759 | ENSG00000126882 | FAM78A | 0,367 | 4,179 | 4,10E-05 | 2,13E-02 |
| cg24224501 | ENSG00000173369 | C1QB | 0,978 | 4,181 | 4,06E-05 | 2,13E-02 |
| cg24224501 | ENSG00000196329 | GIMAP5 | 0,995 | 4,180 | 4,08E-05 | 2,13E-02 |
| cg20372759 | ENSG00000124224 | PPP4R1L | -0,334 | -4,172 | 4,23E-05 | 2,17E-02 |
| cg20372759 | ENSG00000112877 | CEP72 | -0,241 | -4,161 | 4,41E-05 | 2,24E-02 |
| cg24224501 | ENSG00000142185 | TRPM2 | 0,882 | 4,160 | 4,43E-05 | 2,24E-02 |

| | | | | | | |
|---|---|---|---|---|---|---|
| cg20372759 | ENSG00000163221 | S100A12 | 0,928 | 4,142 | 4,77E-05 | 2,38E-02 |
| cg20372759 | ENSG00000163606 | CD200R1 | -0,570 | -4,139 | 4,84E-05 | 2,38E-02 |
| cg24224501 | ENSG00000003137 | CYP26B1 | 1,376 | 4,138 | 4,85E-05 | 2,38E-02 |
| cg24224501 | ENSG00000167286 | CD3D | 0,980 | 4,135 | 4,91E-05 | 2,38E-02 |
| cg24224501 | ENSG00000180096 | SEPT1 | 1,283 | 4,130 | 5,01E-05 | 2,41E-02 |
| cg20372759 | ENSG00000120708 | TGFBI | 0,334 | 4,127 | 5,08E-05 | 2,42E-02 |
| cg01870976 | ENSG00000187094 | CCK | -0,647 | -4,118 | 5,26E-05 | 2,44E-02 |
| cg20372759 | ENSG00000182578 | CSF1R | 0,362 | 4,119 | 5,24E-05 | 2,44E-02 |
| cg20372759 | ENSG00000272931 | | -0,317 | -4,121 | 5,19E-05 | 2,44E-02 |
| cg01870976 | ENSG00000082397 | EPB41L3 | 0,531 | 4,115 | 5,31E-05 | 2,44E-02 |
| cg24224501 | ENSG00000105122 | RASAL3 | 0,762 | 4,113 | 5,37E-05 | 2,44E-02 |
| cg20372759 | ENSG00000182606 | TRAK1 | -0,164 | -4,104 | 5,57E-05 | 2,51E-02 |
| cg24224501 | ENSG00000170500 | LONRF2 | -1,015 | -4,101 | 5,64E-05 | 2,52E-02 |
| cg24224501 | ENSG00000088827 | SIGLEC1 | 1,223 | 4,096 | 5,75E-05 | 2,53E-02 |
| cg24224501 | ENSG00000231389 | HLA-DPA1 | 0,847 | 4,095 | 5,76E-05 | 2,53E-02 |
| cg24224501 | ENSG00000197253 | TPSB2 | -2,831 | -4,081 | 6,11E-05 | 2,66E-02 |
| cg01870976 | ENSG00000130635 | COL5A1 | 0,679 | 4,071 | 6,36E-05 | 2,74E-02 |
| cg20372759 | ENSG00000187094 | CCK | -0,596 | -4,069 | 6,42E-05 | 2,74E-02 |
| cg24224501 | ENSG00000175899 | A2M | 0,719 | 4,066 | 6,49E-05 | 2,74E-02 |
| cg24224501 | ENSG00000211772 | TRBC2 | 0,943 | 4,065 | 6,50E-05 | 2,74E-02 |
| cg01870976 | ENSG00000179776 | CDH5 | 1,008 | 4,059 | 6,67E-05 | 2,79E-02 |
| cg20372759 | ENSG00000108950 | FAM20A | 0,377 | 4,055 | 6,77E-05 | 2,81E-02 |
| cg20372759 | ENSG00000185022 | MAFF | 0,379 | 4,045 | 7,04E-05 | 2,88E-02 |
| cg24224501 | ENSG00000211459 | MT-RNR1 | -0,702 | -4,046 | 7,02E-05 | 2,88E-02 |
| cg24224501 | ENSG00000264198 | | 0,997 | 4,034 | 7,36E-05 | 2,98E-02 |
| cg20372759 | ENSG00000137101 | CD72 | 0,342 | 4,026 | 7,60E-05 | 3,06E-02 |
| cg24224501 | ENSG00000179583 | CIITA | 0,778 | 4,020 | 7,80E-05 | 3,11E-02 |
| cg24224501 | ENSG00000105374 | NKG7 | 1,216 | 4,015 | 7,93E-05 | 3,14E-02 |
| cg20372759 | ENSG00000168995 | SIGLEC7 | 0,721 | 4,008 | 8,18E-05 | 3,16E-02 |
| cg20372759 | ENSG00000179776 | CDH5 | 0,929 | 4,011 | 8,09E-05 | 3,16E-02 |
| cg24224501 | ENSG00000182578 | CSF1R | 0,741 | 4,009 | 8,13E-05 | 3,16E-02 |
| cg20372759 | ENSG00000163734 | CXCL3 | 0,666 | 4,001 | 8,42E-05 | 3,21E-02 |
| cg24224501 | ENSG00000137078 | SIT1 | 1,185 | 4,000 | 8,43E-05 | 3,21E-02 |
| cg20372759 | ENSG00000116299 | KIAA1324 | -0,255 | -3,995 | 8,60E-05 | 3,23E-02 |
| cg20372759 | ENSG00000135063 | FAM189A2 | -0,265 | -3,995 | 8,59E-05 | 3,23E-02 |
| cg24224501 | ENSG00000184371 | CSF1 | 0,725 | 3,990 | 8,76E-05 | 3,27E-02 |
| cg20372759 | ENSG00000131620 | ANO1 | -0,452 | -3,988 | 8,83E-05 | 3,27E-02 |
| cg01870976 | ENSG00000012660 | ELOVL5 | -0,259 | -3,982 | 9,07E-05 | 3,32E-02 |
| cg24224501 | ENSG00000126882 | FAM78A | 0,738 | 3,981 | 9,11E-05 | 3,32E-02 |
| cg20372759 | ENSG00000163399 | ATP1A1 | -0,136 | -3,972 | 9,44E-05 | 3,40E-02 |
| cg24224501 | ENSG00000106991 | ENG | 0,635 | 3,970 | 9,51E-05 | 3,40E-02 |
| cg24224501 | ENSG00000108798 | ABI3 | 0,831 | 3,970 | 9,51E-05 | 3,40E-02 |
| cg24224501 | ENSG00000198886 | MT-ND4 | -0,545 | -3,967 | 9,62E-05 | 3,42E-02 |
| cg01870976 | ENSG00000092964 | DPYSL2 | 0,317 | 3,955 | 1,01E-04 | 3,55E-02 |
| cg24224501 | ENSG00000125037 | EMC3 | -0,246 | -3,940 | 1,07E-04 | 3,71E-02 |
| cg24224501 | ENSG00000136541 | ERMN | -1,548 | -3,941 | 1,06E-04 | 3,71E-02 |
| cg01870976 | ENSG00000107960 | OBFC1 | -0,156 | -3,938 | 1,08E-04 | 3,71E-02 |
| cg20372759 | ENSG00000186818 | LILRB4 | 0,420 | 3,937 | 1,08E-04 | 3,71E-02 |
| cg24224501 | ENSG00000081059 | TCF7 | 0,554 | 3,918 | 1,17E-04 | 3,98E-02 |
| cg01870976 | ENSG00000142185 | TRPM2 | 0,425 | 3,909 | 1,21E-04 | 4,08E-02 |
| cg24224501 | ENSG00000100628 | ASB2 | 0,909 | 3,905 | 1,22E-04 | 4,12E-02 |
| cg24224501 | ENSG00000137094 | DNAJB5 | 0,630 | 3,901 | 1,24E-04 | 4,16E-02 |

| | | | | | | |
|---|---|---|---|---|---|---|
| cg20372759 | ENSG00000175356 | SCUBE2 | -0,548 | -3,896 | 1,27E-04 | 4,21E-02 |
| cg20372759 | ENSG00000076864 | RAP1GAP | -0,358 | -3,890 | 1,30E-04 | 4,29E-02 |
| cg24224501 | ENSG00000198712 | MT-CO2 | -0,557 | -3,886 | 1,32E-04 | 4,32E-02 |
| cg20372759 | ENSG00000130635 | COL5A1 | 0,605 | 3,884 | 1,33E-04 | 4,33E-02 |
| cg01870976 | ENSG00000109861 | CTSC | -0,216 | -3,874 | 1,38E-04 | 4,38E-02 |
| cg01870976 | ENSG00000264456 | | 0,566 | 3,876 | 1,37E-04 | 4,38E-02 |
| cg20372759 | ENSG00000092964 | DPYSL2 | 0,290 | 3,877 | 1,37E-04 | 4,38E-02 |
| cg24224501 | ENSG00000161791 | FMNL3 | 0,667 | 3,873 | 1,39E-04 | 4,38E-02 |
| cg24224501 | ENSG00000186810 | CXCR3 | 1,124 | 3,878 | 1,36E-04 | 4,38E-02 |
| cg20372759 | ENSG00000072694 | FCGR2B | 0,459 | 3,869 | 1,41E-04 | 4,43E-02 |
| cg01870976 | ENSG00000088827 | SIGLEC1 | 0,591 | 3,865 | 1,43E-04 | 4,44E-02 |
| cg24224501 | ENSG00000187527 | ATP13A5 | -0,676 | -3,865 | 1,43E-04 | 4,44E-02 |
| cg01870976 | ENSG00000170500 | LONRF2 | -0,489 | -3,857 | 1,48E-04 | 4,55E-02 |
| cg20372759 | ENSG00000163735 | CXCL5 | 1,126 | 3,855 | 1,48E-04 | 4,55E-02 |
| cg01870976 | ENSG00000272931 | | -0,319 | -3,852 | 1,50E-04 | 4,58E-02 |
| cg20372759 | ENSG00000107562 | CXCL12 | 0,808 | 3,843 | 1,56E-04 | 4,69E-02 |
| cg20372759 | ENSG00000155850 | SLC26A2 | -0,363 | -3,843 | 1,55E-04 | 4,69E-02 |
| cg24224501 | ENSG00000156234 | CXCL13 | 1,909 | 3,839 | 1,58E-04 | 4,72E-02 |
| cg20372759 | ENSG00000187474 | FPR3 | 0,458 | 3,831 | 1,63E-04 | 4,73E-02 |
| cg24224501 | ENSG00000061337 | LZTS1 | 1,518 | 3,832 | 1,62E-04 | 4,73E-02 |
| cg24224501 | ENSG00000140464 | PML | 0,564 | 3,832 | 1,62E-04 | 4,73E-02 |
| cg24224501 | ENSG00000196440 | ARMCX4 | -0,754 | -3,832 | 1,62E-04 | 4,73E-02 |
| cg24224501 | ENSG00000237943 | PRKCQ-AS1 | 1,285 | 3,833 | 1,62E-04 | 4,73E-02 |
| cg01870976 | ENSG00000137101 | CD72 | 0,349 | 3,820 | 1,70E-04 | 4,87E-02 |
| cg20372759 | ENSG00000265206 | MIR142 | 0,524 | 3,820 | 1,70E-04 | 4,87E-02 |
| cg24224501 | ENSG00000083457 | ITGAE | 0,359 | 3,819 | 1,70E-04 | 4,87E-02 |
| cg20372759 | ENSG00000163220 | S100A9 | 0,484 | 3,812 | 1,75E-04 | 4,97E-02 |
| cg20372759 | ENSG00000163053 | SLC16A14 | -0,350 | -3,811 | 1,76E-04 | 4,98E-02 |

Table S8. Gene modules identified from eQTM genes by WGCNA (weighted correlation network analysis)

| Module1 | Module2 |
|---|---|
| ZAP70 | SIGLEC1 |
| FAM78A | CD72 |
| ANO1 | TRPM2 |
| CD36 | GPR68 |
| NMUR1 | TGFBI |
| FCRL6 | CSF1R |
| TMC8 | LILRB4 |
| CD6 | TMEM176B |
| TCF7 | ENG |
| ITGAE | SLCO2B1 |
| IL2RB | C1QC |
| ASB2 | C1QB |
| RASAL3 | |
| NKG7 | |
| SPOCK2 | |
| ABI3 | |
| CD5 | |
| CD2 | |
| ACY3 | |
| SIT1 | |
| PML | |
| FGD2 | |
| CD8A | |
| CCL5 | |
| FMNL3 | |
| SDC3 | |
| TMEM71 | |
| CD3D | |
| MSC | |
| CIITA | |
| SEPT1 | |
| PRF1 | |
| LCK | |
| CSF1 | |
| CXCR3 | |
| PDCD1 | |
| GIMAP5 | |
| GZMM | |
| CD3E | |
| HLA-DOA | |
| TRBC2 | |
| TRAC | |
| HLA-DPA1 | |
| ENSG00000264198 | |

Table S9 Sample characteristics of single-cell RNASeq dataset

| | | N |
|---|---|---|
| Total | | 9 |
| Age | | 51.4 ± 8.4 |
| Gender male (%) | | 6 (66.7%) |
| BMI | | 28.1 ± 4.9 |
| Smoking (%) | | 2 (22.2%) |
| Disease | | |
| | Asthma (%) | 4 (44.4%) |
| | Rhinitis (%) | 3 (33.3%) |
| | Eczema (%) | 3 (33.3%) |

Table S10. MeQTL analysis of three CpG sites with allergy-related SNPs identified by Ferreira et al. (2017)

| SNP ID | CHROM | effect_allele | other_allele | risk allele of allergic disease | POS | Gene | CpG ID | beta | se | pvalue |
|---|---|---|---|---|---|---|---|---|---|---|
| rs9372120 | 6 | G | T | G | 106667535 | [ATG5] | cg20372759 | -0,203 | 0,060 | 8,61E-04 |
| rs144829310 | 9 | T | G | T | 6208030 | RANBP6--[]-IL33 | cg24224501 | 0,068 | 0,026 | 9,28E-03 |
| rs7130753 | 11 | T | C | C | 111470567 | LAYN-[]-SIK2 | cg24224501 | 0,049 | 0,021 | 0,019 |
| rs9372120 | 6 | G | T | G | 106667535 | [ATG5] | cg01870976 | -0,121 | 0,052 | 0,022 |
| rs3749833 | 5 | C | T | C | 131799626 | [C5orf56] | cg01870976 | -0,091 | 0,039 | 0,022 |
| rs76167968 | 1 | C | T | T | 35681738 | SFPQ-[]-ZMYM4 | cg24224501 | 0,082 | 0,036 | 0,023 |
| rs2104047 | 14 | C | T | T | 68754417 | [RAD51B] | cg01870976 | -0,087 | 0,038 | 0,023 |
| rs10486391 | 7 | G | A | A | 20376018 | [ITGB8] | cg20372759 | -0,084 | 0,037 | 0,024 |
| rs4574025 | 18 | T | C | T | 60009814 | [TNFRSF11A] | cg20372759 | -0,087 | 0,039 | 0,027 |
| rs56129466 | 11 | G | A | A | 128158189 | KIRREL3-AS3---[]--ETS1 | cg24224501 | -0,047 | 0,022 | 0,031 |
| rs11204896 | 1 | G | C | C | 151796742 | [RORC] | cg01870976 | 0,125 | 0,058 | 0,032 |
| rs6869502 | 5 | T | A | T | 110166083 | SLC25A46-[]--TSLP | cg20372759 | -0,119 | 0,058 | 0,043 |
| rs4848612 | 2 | A | G | A | 112388538 | BCL2L11--[]--ANAPC1 | cg24224501 | -0,039 | 0,019 | 0,043 |
| rs3749833 | 5 | C | T | C | 131799626 | [C5orf56] | cg20372759 | -0,091 | 0,046 | 0,047 |
| rs5029937 | 6 | T | G | G | 138195151 | [TNFAIP3] | cg24224501 | -0,101 | 0,051 | 0,047 |

Gene: the nearest genes of the SNP

Ferreira, M. A. *et al.* Shared genetic origin of asthma, hay fever and eczema elucidates allergic disease biology. *Nat. Genet.* **49**, 1752–1757 (2017)

Table S11. Mediation analysis of SNP (rs9372120) - DNA methylation (cg20372759) - allergic disease

| Model | variable tested | Estmate | SE | t value | P value |
|---|---|---|---|---|---|
| 1. Allergic disese ~ SNP | SNP | 0,099 | 0,043 | 2,297 | 0,022 |
| 2. DNAm ~ SNP | SNP | -0,229 | 0,06 | -3,849 | 0,00013 |
| 3. Alleri disease ~ SNP +DNAm | SNP | 0,024 | 0,039 | 0,615 | 0,539 |
| Mediation analysis | | Estimate | 95% CI Lower | 95% CI Upper | P value |
| | ACME | 0,075 | 0,036 | 0,12 | <2E-16 |
| | ADE | 0,024 | -0,058 | 0,11 | 0,554 |
| | Total Effect | 0,099 | 0,016 | 0,19 | 0,018 |
| | Proportion Mediated | 0,757 | 0,327 | 3,49 | 0,018 |

ACME: Average Causal Mediation Effects

ADE: Average Direct Effects

Table S12. Sample characteristics of 16-year follow-up on subjects (included/ not included in the analyses), and subjects who did not participate in the 16-year follow-up medical examination

| | | Subjects who participated in 16-year follow-up medical examination | | Subjects who did not participate in the 16-year follow-up medical examination (n=3161) |
|---|---|---|---|---|
| | | Subjects in the analyses (n=348) | Subjects not in the analyses (n=454) | |
| General information | Age (years) | 16.3 ± 0.2 | 16.4 ± 0.2 | NA |
| | Sex: number of males | 169 (48.6%) | 217 (47.8%) | 1668 (52.8%) * |
| Baseline information | Atopic mother | 122/ 348 (35.1%) | 135/ 454 (29.7%) | 980/ 3161 (31.0%) |
| | Atopic father | 112/ 348 (32.2%) | 154/ 453 (34.0%) | 951/ 3156 (30.1%) |
| | Breast feeding | 305/ 348 (87.6%) | 400/ 454 (88.1%) | 2495/ 3094 (80.6%)* |
| | Maternal education | | | * |
| | Low | 56/ 348 (16.1%) | 65/ 454 (14.3%) | 773/ 3005 (25.7%) |
| | Medium | 135/ 348 (38.8%) | 181/ 454 (39.9%) | 1266/ 3005 (42.1%) |
| | High | 157/ 348 (45.1%) | 208/ 454 (45.8%) | 966/ 3005 (32.1%) |
| | Maternal smoking | 54/ 348 (15.5%) | 51/ 447 (11.4%) | 598/ 3105 (19.3%) * |
| | Birth weight (g) | 3564.4 ± 533.9 | 3547.5 ± 507.9 | 3495.0 ± 552.3 * |
| Information of 16 years follow-up | Asthma | 23/ 348 (6.6%) | 43/ 418 (10.3 %) | |
| | Rhinitis | 59/ 348 (17.0%) | 74/ 409 (18.1%) | |
| | Eczema | 29/ 348 (8.3%) | **56/ 419 (13.4%)** | |
| | Positive allergen-specific IgE | 162/ 348 (46.6%) | 182/ 373 (48.8%) | |
| | Allergy | 67/ 348 (19.3%) | 93/ 407 (22.9%) | |
| | Pets | 183/ 291 (62.9%) | **206/ 375 (54.9%)** | |
| | Current smoking | 37/ 297 (12.5%) | 47/ 356 (13.2%) | |

Breast feeding: includes any breast feeding

Maternal smoking: maternal smoking during at least the first 4 weeks of pregnancy

Continuous variables are presented as mean ± SD; categorical variables are presented as number of ("yes")/ (total number (non-missing)) (percentage)

**Bold**: significant difference compared to subjects in the analyses ($P<0.05$), t test for continous variable and chi-square test for categorical variable

* significant difference compared to subjects who participated in the 16-year follow-up medical examination ($P<0.05$), t test for continous variable and chi-square test for categorical variable

Table S13. Overview of all used predictive features ordered by data layer

| Data layer | Feature name |
|---|---|
| Host | age |
| Host | gender |
| Environment | older_siblings_home |
| Environment | cats_dogs_during_pregnancy |
| Environment | smoking_mother_pregnancy |
| Perinatal | low_birth_weight |
| Perinatal | any_breastfeeding |
| Genetics | Allergy_PRS |
| Genetics | Asthma_PRS |
| Genetics | Sensitization_PRS |
| Genetics | Eczema_PRS |
| Genetics | Rhinitis_PRS |
| Genetics | SNP.1.2510755 |
| Genetics | SNP.1.8482078 |
| Genetics | SNP.1.25251923 |
| Genetics | SNP.1.35681738 |
| Genetics | SNP.1.151796742 |
| Genetics | SNP.1.154426970 |
| Genetics | SNP.1.161185058 |
| Genetics | SNP.1.167431352 |
| Genetics | SNP.1.173146921 |
| Genetics | SNP.1.226914734 |
| Genetics | SNP.2.8442248 |
| Genetics | SNP.2.102926362 |
| Genetics | SNP.2.102941311 |
| Genetics | SNP.2.112388538 |
| Genetics | SNP.2.113590467 |
| Genetics | SNP.2.143831599 |
| Genetics | SNP.2.198950240 |
| Genetics | SNP.2.228707862 |
| Genetics | SNP.20.62322699 |
| Genetics | SNP.21.36467830 |
| Genetics | SNP.3.33069091 |
| Genetics | SNP.3.72394852 |
| Genetics | SNP.3.101242751 |
| Genetics | SNP.3.187633268 |
| Genetics | SNP.3.187793833 |
| Genetics | SNP.3.188133336 |
| Genetics | SNP.3.188402586 |
| Genetics | SNP.4.4775401 |
| Genetics | SNP.4.38798648 |
| Genetics | SNP.4.123316076 |
| Genetics | SNP.5.14610309 |
| Genetics | SNP.5.35862841 |
| Genetics | SNP.5.40492655 |
| Genetics | SNP.5.110166083 |
| Genetics | SNP.5.110401872 |
| Genetics | SNP.5.118684297 |
| Genetics | SNP.5.131799626 |
| Genetics | SNP.5.131989136 |
| Genetics | SNP.5.131996500 |
| Genetics | SNP.5.140925362 |

| | |
|---|---|
| Genetics | SNP.5.141494934 |
| Genetics | SNP.5.176782218 |
| Genetics | SNP.6.31323012 |
| Genetics | SNP.6.31351664 |
| Genetics | SNP.6.31574525 |
| Genetics | SNP.6.33046752 |
| Genetics | SNP.6.33647058 |
| Genetics | SNP.6.90987512 |
| Genetics | SNP.6.106667535 |
| Genetics | SNP.6.128294709 |
| Genetics | SNP.6.138195151 |
| Genetics | SNP.6.157419508 |
| Genetics | SNP.7.20376018 |
| Genetics | SNP.7.20560996 |
| Genetics | SNP.7.28156887 |
| Genetics | SNP.7.50253897 |
| Genetics | SNP.7.77018542 |
| Genetics | SNP.8.128814091 |
| Genetics | SNP.9.5064193 |
| Genetics | SNP.9.6051399 |
| Genetics | SNP.9.6208030 |
| Genetics | SNP.9.123650534 |
| Genetics | SNP.10.6074451 |
| Genetics | SNP.10.6094697 |
| Genetics | SNP.10.8841669 |
| Genetics | SNP.10.8936162 |
| Genetics | SNP.10.9049253 |
| Genetics | SNP.10.9064361 |
| Genetics | SNP.10.64382359 |
| Genetics | SNP.10.104225832 |
| Genetics | SNP.11.65551957 |
| Genetics | SNP.11.76293758 |
| Genetics | SNP.11.76299431 |
| Genetics | SNP.11.76343428 |
| Genetics | SNP.11.95425526 |
| Genetics | SNP.11.111470567 |
| Genetics | SNP.11.118743286 |
| Genetics | SNP.11.128158189 |
| Genetics | SNP.12.48196982 |
| Genetics | SNP.12.50345671 |
| Genetics | SNP.12.56401085 |
| Genetics | SNP.12.57489709 |
| Genetics | SNP.12.111932800 |
| Genetics | SNP.12.121363724 |
| Genetics | SNP.13.41173408 |
| Genetics | SNP.13.73627275 |
| Genetics | SNP.14.35761675 |
| Genetics | SNP.14.68754417 |
| Genetics | SNP.14.75968608 |
| Genetics | SNP.14.103235012 |
| Genetics | SNP.15.61068347 |
| Genetics | SNP.15.67448363 |
| Genetics | SNP.16.11277358 |
| Genetics | SNP.16.11491007 |

| | |
|---|---|
| Genetics | SNP.17.38069076 |
| Genetics | SNP.17.38149033 |
| Genetics | SNP.17.43430696 |
| Genetics | SNP.17.47398070 |
| Genetics | SNP.18.52336175 |
| Genetics | SNP.18.60009814 |
| Genetics | SNP.19.33721455 |
| Blood DNA methylation | cg00318756 |
| Blood DNA methylation | cg00148881 |
| Blood DNA methylation | cg00251536 |
| Blood DNA methylation | cg00321867 |
| Blood DNA methylation | cg00099427 |
| Blood DNA methylation | cg00001583 |
| Blood DNA methylation | cg00030466 |
| Blood DNA methylation | cg00309462 |
| Blood DNA methylation | cg00298020 |
| Blood DNA methylation | cg00065905 |
| Blood DNA methylation | cg00152577 |
| Blood DNA methylation | cg00300039 |
| Blood DNA methylation | cg00081019 |
| Blood DNA methylation | cg00297950 |
| Blood DNA methylation | cg00035237 |
| Blood DNA methylation | cg00177390 |
| Blood DNA methylation | cg00042144 |
| Blood DNA methylation | cg00346501 |
| Blood DNA methylation | cg00027570 |
| Blood DNA methylation | cg00152515 |
| Blood DNA methylation | cg00304520 |
| Blood DNA methylation | cg00248795 |
| Blood DNA methylation | cg00254532 |
| Blood DNA methylation | cg00055434 |
| Blood DNA methylation | cg00252804 |
| Blood DNA methylation | cg00107970 |
| Blood DNA methylation | cg00053086 |
| Blood DNA methylation | cg00155429 |
| Blood DNA methylation | cg00090648 |
| Blood DNA methylation | cg00311307 |
| Blood DNA methylation | cg00040367 |
| Blood DNA methylation | cg00097626 |
| Blood DNA methylation | cg00382944 |
| Blood DNA methylation | cg00347677 |
| Blood DNA methylation | cg00081799 |
| Blood DNA methylation | cg00009916 |
| Blood DNA methylation | cg00090103 |
| Blood DNA methylation | cg00036263 |
| Blood DNA methylation | cg00035249 |
| Blood DNA methylation | cg00034416 |
| Blood DNA methylation | cg00153856 |
| Blood DNA methylation | cg00062020 |
| Blood DNA methylation | cg00097228 |
| Blood DNA methylation | cg00049400 |
| Blood DNA methylation | cg00254258 |
| Blood DNA methylation | cg00021933 |
| Blood DNA methylation | cg00154064 |

| | |
|---|---|
| Blood DNA methylation | cg00164462 |
| Blood DNA methylation | cg00106446 |
| Blood DNA methylation | cg00296458 |
| Blood DNA methylation | cg00309178 |
| Blood DNA methylation | cg00297432 |
| Blood DNA methylation | cg00311883 |
| Blood DNA methylation | cg00091162 |
| Blood DNA methylation | cg00003202 |
| Blood DNA methylation | cg00046018 |
| Blood DNA methylation | cg00021123 |
| Blood DNA methylation | cg00173405 |
| Blood DNA methylation | cg00068377 |
| Blood DNA methylation | cg00116658 |
| Blood DNA methylation | cg00349061 |
| Blood DNA methylation | cg00042657 |
| Blood DNA methylation | cg00089372 |
| Blood DNA methylation | cg00042356 |
| Blood DNA methylation | cg00116430 |
| Blood DNA methylation | cg00290506 |
| Blood DNA methylation | cg00002719 |
| Blood DNA methylation | cg00329748 |
| Blood DNA methylation | cg00305774 |
| Blood DNA methylation | cg00001349 |
| Blood DNA methylation | cg00305374 |
| Blood DNA methylation | cg00357368 |
| Blood DNA methylation | cg00305285 |
| Blood DNA methylation | cg00093900 |
| Blood DNA methylation | cg00251250 |
| Blood DNA methylation | cg00150520 |
| Blood DNA methylation | cg00145911 |
| Blood DNA methylation | cg00000957 |
| Blood DNA methylation | cg00151906 |
| Blood DNA methylation | cg00170536 |
| Blood DNA methylation | cg00090197 |
| Blood DNA methylation | cg00339300 |
| Blood DNA methylation | cg00026222 |
| Blood DNA methylation | cg00300879 |
| Blood DNA methylation | cg00002837 |
| Blood DNA methylation | cg00015930 |
| Blood DNA methylation | cg00023492 |
| Blood DNA methylation | cg00031063 |
| Blood DNA methylation | cg00046625 |
| Blood DNA methylation | cg00052482 |
| Blood DNA methylation | cg00060374 |
| Blood DNA methylation | cg00060606 |
| Blood DNA methylation | cg00068412 |
| Blood DNA methylation | cg00095594 |
| Blood DNA methylation | cg00102714 |
| Blood DNA methylation | cg00125159 |
| Blood DNA methylation | cg00149684 |
| Blood DNA methylation | cg00163784 |
| Blood DNA methylation | cg00166706 |
| Blood DNA methylation | cg00172597 |
| Blood DNA methylation | cg00172812 |

| | |
|---|---|
| Blood DNA methylation | cg00249030 |
| Blood DNA methylation | cg00252701 |
| Blood DNA methylation | cg00253185 |
| Blood DNA methylation | cg00257659 |
| Blood DNA methylation | cg00295572 |
| Blood DNA methylation | cg00306390 |
| Blood DNA methylation | cg00346208 |
| Blood DNA methylation | cg00349607 |
| Blood DNA methylation | cg00371107 |
| Blood DNA methylation | cg00383735 |
| Blood DNA methylation | cg00172408 |
| Blood DNA methylation | cg00155063 |
| Blood DNA methylation | cg00115101 |
| Blood DNA methylation | cg00087735 |
| Blood DNA methylation | cg00011717 |
| Blood DNA methylation | cg00009292 |
| Blood DNA methylation | cg00107241 |
| Blood DNA methylation | cg00160667 |
| Blood DNA methylation | cg00016238 |
| Blood DNA methylation | cg00064255 |
| Blood DNA methylation | cg00113194 |
| Blood DNA methylation | cg00049718 |
| Blood DNA methylation | cg00165011 |
| Blood DNA methylation | cg00097573 |
| Blood DNA methylation | cg00007898 |
| Blood DNA methylation | cg00115313 |
| Blood DNA methylation | cg00161970 |
| Blood DNA methylation | cg00083652 |
| Blood DNA methylation | cg00255821 |
| Blood DNA methylation | cg00342358 |
| Blood DNA methylation | cg00066671 |
| Blood DNA methylation | cg00044463 |
| Blood DNA methylation | cg00038675 |
| Blood DNA methylation | cg00149537 |
| Blood DNA methylation | cg00086266 |
| Blood DNA methylation | cg00106073 |
| Blood DNA methylation | cg00031456 |
| Blood DNA methylation | cg00058299 |
| Blood DNA methylation | cg00034101 |
| Blood DNA methylation | cg00114008 |
| Blood DNA methylation | cg00172603 |
| Blood DNA methylation | cg00082497 |
| Blood DNA methylation | cg00114029 |
| Blood DNA methylation | cg00007036 |
| Blood DNA methylation | cg00347563 |
| Blood DNA methylation | cg00176672 |
| Blood DNA methylation | cg00040446 |
| Blood DNA methylation | cg00337821 |
| Blood DNA methylation | cg00256155 |
| Blood DNA methylation | cg00256767 |
| Blood DNA methylation | cg00124011 |
| Blood DNA methylation | cg00099216 |
| Blood DNA methylation | cg00021576 |
| Blood DNA methylation | cg00069860 |

| | |
|---|---|
| Blood DNA methylation | cg00173787 |
| Blood DNA methylation | cg00290994 |
| Blood DNA methylation | cg00002028 |
| Blood DNA methylation | cg00341060 |
| Blood DNA methylation | cg00016996 |
| Blood DNA methylation | cg00329697 |
| Blood DNA methylation | cg00296079 |
| Blood DNA methylation | cg00371368 |
| Blood DNA methylation | cg00328972 |
| Blood DNA methylation | cg00350519 |
| Blood DNA methylation | cg00003287 |
| Blood DNA methylation | cg00249522 |
| Blood DNA methylation | cg00325919 |
| Blood DNA methylation | cg00063077 |
| Blood DNA methylation | cg00112780 |
| Blood DNA methylation | cg00302521 |
| Blood DNA methylation | cg00287370 |
| Blood DNA methylation | cg00385863 |
| Blood DNA methylation | cg00008004 |
| Blood DNA methylation | cg00101986 |
| Blood DNA methylation | cg00255925 |
| Blood DNA methylation | cg00043324 |
| Blood DNA methylation | cg00306532 |
| Blood DNA methylation | cg00118468 |
| Blood DNA methylation | cg00061769 |
| Blood DNA methylation | cg00037072 |
| Blood DNA methylation | cg00352560 |
| Blood DNA methylation | cg00169776 |
| Blood DNA methylation | cg00257786 |
| Blood DNA methylation | cg00346985 |
| Blood DNA methylation | cg00345792 |
| Blood DNA methylation | cg00329300 |
| Blood DNA methylation | cg00354179 |
| Blood DNA methylation | cg00151676 |
| Blood DNA methylation | cg00044796 |
| Blood DNA methylation | cg00299820 |
| Blood DNA methylation | cg00298532 |
| Blood DNA methylation | cg00173141 |
| Blood DNA methylation | cg00359395 |
| Blood DNA methylation | cg00069017 |
| Blood DNA methylation | cg00350652 |
| Blood DNA methylation | cg00157179 |
| Blood DNA methylation | cg00318766 |
| Blood DNA methylation | cg00154982 |
| Blood DNA methylation | cg00114084 |
| Blood DNA methylation | cg00091285 |
| Blood DNA methylation | cg00085403 |
| Blood DNA methylation | cg00152008 |
| Blood DNA methylation | cg00383290 |
| Blood DNA methylation | cg00171113 |
| Blood DNA methylation | cg00305874 |
| Blood DNA methylation | cg00171783 |
| Blood DNA methylation | cg00344178 |
| Blood DNA methylation | cg00172330 |

| | |
|---|---|
| Blood DNA methylation | cg00151709 |
| Blood DNA methylation | cg00373131 |
| Blood DNA methylation | cg00306607 |
| Blood DNA methylation | cg00008647 |
| Blood DNA methylation | cg00066499 |
| Blood DNA methylation | cg00065215 |
| Blood DNA methylation | cg00355315 |
| Blood DNA methylation | cg00310968 |
| Blood DNA methylation | cg00228891 |
| Blood DNA methylation | cg00105470 |
| Nasal DNA methylation | cg20372759 |
| Nasal DNA methylation | cg01870976 |
| Nasal DNA methylation | cg24224501 |
| Nasal DNA methylation | cg22862094 |
| Nasal DNA methylation | cg16027132 |
| Nasal DNA methylation | cg20790648 |
| Nasal DNA methylation | cg24707200 |
| Nasal DNA methylation | cg15006973 |
| Nasal DNA methylation | cg08844313 |
| Nasal DNA methylation | cg22689016 |
| Nasal DNA methylation | cg00285620 |
| Nasal DNA methylation | cg03387497 |
| Nasal DNA methylation | cg08341667 |
| Nasal DNA methylation | cg25020944 |
| Nasal DNA methylation | cg02333649 |
| Nasal DNA methylation | cg03565274 |
| Nasal DNA methylation | cg18749617 |
| Nasal DNA methylation | cg23387401 |
| Nasal DNA methylation | cg01280881 |
| Nasal DNA methylation | cg20337028 |
| Nasal DNA methylation | cg11058904 |
| Nasal DNA methylation | cg19610615 |
| Nasal DNA methylation | cg10549071 |
| Nasal DNA methylation | cg17223698 |
| Nasal DNA methylation | cg15294341 |
| Nasal DNA methylation | cg03668556 |
| Nasal DNA methylation | cg00664723 |
| Nasal DNA methylation | cg13586696 |
| Nasal DNA methylation | cg01859321 |
| Nasal DNA methylation | cg02702515 |
| Nasal DNA methylation | cg02648939 |
| Nasal DNA methylation | cg03875819 |
| Nasal DNA methylation | cg05598678 |
| Nasal DNA methylation | cg08167132 |
| Nasal DNA methylation | cg07686035 |
| Nasal DNA methylation | cg08097877 |
| Nasal DNA methylation | cg22855021 |
| Nasal DNA methylation | cg10099207 |
| Nasal DNA methylation | cg01062020 |
| Nasal DNA methylation | cg27073349 |
| Nasal DNA methylation | cg14232889 |
| Nasal DNA methylation | cg23005227 |
| Nasal DNA methylation | cg10054641 |
| Nasal DNA methylation | cg08175352 |

| | |
|---|---|
| Nasal DNA methylation | cg09562938 |
| Nasal DNA methylation | cg10830021 |
| Nasal DNA methylation | cg12805346 |
| Nasal DNA methylation | cg09649586 |
| Nasal DNA methylation | cg06255006 |
| Nasal DNA methylation | cg16118839 |
| Nasal DNA methylation | cg09997532 |
| Nasal DNA methylation | cg18297196 |
| Nasal DNA methylation | cg10583485 |
| Nasal DNA methylation | cg06419562 |
| Nasal DNA methylation | cg12716639 |
| Nasal DNA methylation | cg14588406 |
| Nasal DNA methylation | cg09171565 |
| Nasal DNA methylation | cg00049323 |
| Nasal DNA methylation | cg01634388 |
| Nasal DNA methylation | cg07178994 |
| Nasal DNA methylation | cg15028507 |
| Nasal DNA methylation | cg00970361 |
| Nasal DNA methylation | cg01445399 |
| Nasal DNA methylation | cg24975342 |
| Nasal DNA methylation | cg09327821 |
| Nasal DNA methylation | cg00146864 |
| Nasal DNA methylation | cg04891688 |
| Nasal DNA methylation | cg09285543 |
| Nasal DNA methylation | cg15388974 |
| Nasal DNA methylation | cg13961587 |
| Nasal DNA methylation | cg03794679 |
| Nasal DNA methylation | cg18894440 |
| Nasal DNA methylation | cg12940991 |
| Nasal DNA methylation | cg07094693 |
| Nasal DNA methylation | cg08816988 |
| Nasal DNA methylation | cg04120420 |
| Nasal DNA methylation | cg10305969 |
| Nasal DNA methylation | cg08121164 |
| Nasal DNA methylation | cg26793720 |
| Nasal DNA methylation | cg00513564 |
| Nasal DNA methylation | cg02912316 |
| Nasal DNA methylation | cg18291443 |
| Nasal DNA methylation | cg09407660 |
| Nasal DNA methylation | cg16983282 |
| Nasal DNA methylation | cg18504632 |
| Nasal DNA methylation | cg26503079 |
| Nasal DNA methylation | cg14205169 |
| Nasal DNA methylation | cg12147622 |
| Nasal DNA methylation | cg11441533 |
| Nasal DNA methylation | cg21540530 |
| Nasal DNA methylation | cg05659947 |
| Nasal DNA methylation | cg11303839 |
| Nasal DNA methylation | cg10921599 |
| Nasal DNA methylation | cg08776942 |
| Nasal DNA methylation | cg18070053 |
| Nasal DNA methylation | cg21531685 |
| Nasal DNA methylation | cg26438325 |
| Nasal DNA methylation | cg09482780 |

| | |
|---|---|
| Nasal DNA methylation | cg09472600 |
| Nasal DNA methylation | cg22839180 |
| Nasal DNA methylation | cg17863312 |
| Nasal DNA methylation | cg00779056 |
| Nasal DNA methylation | cg06035708 |
| Nasal DNA methylation | cg27208079 |
| Nasal DNA methylation | cg26182317 |
| Nasal DNA methylation | cg00195561 |
| Nasal DNA methylation | cg14299508 |
| Nasal DNA methylation | cg08330217 |
| Nasal DNA methylation | cg16796127 |
| Nasal DNA methylation | cg16177250 |
| Nasal DNA methylation | cg12692069 |
| Nasal DNA methylation | cg19238840 |
| Nasal DNA methylation | cg20511797 |
| Nasal DNA methylation | cg00437411 |
| Nasal DNA methylation | cg14945937 |
| Nasal DNA methylation | cg00764093 |
| Nasal DNA methylation | cg25597055 |
| Nasal DNA methylation | cg14914238 |
| Nasal DNA methylation | cg21291385 |
| Nasal DNA methylation | cg07239613 |
| Nasal DNA methylation | cg16683073 |
| Nasal DNA methylation | cg06675531 |
| Nasal DNA methylation | cg04206484 |
| Nasal DNA methylation | cg14525390 |
| Nasal DNA methylation | cg12875548 |
| Nasal DNA methylation | cg16672659 |
| Nasal DNA methylation | cg27058763 |
| Nasal DNA methylation | cg26656751 |
| Nasal DNA methylation | cg14409166 |
| Nasal DNA methylation | cg09950920 |
| Nasal DNA methylation | cg10719920 |
| Nasal DNA methylation | cg20293725 |
| Nasal DNA methylation | cg07703756 |
| Nasal DNA methylation | cg09362969 |

SNPs annotated as <CHROM>.<POS> using genome build hg19
